# A dynamic microRNA profile that tracks a chemotherapy resistance phenotype in osteosarcoma. Implications for novel therapeutics

**DOI:** 10.1101/2024.06.19.24309087

**Authors:** Christopher E. Lietz, Erik T. Newman, Andrew D. Kelly, David H. Xiang, Ziying Zhang, Nikhil Ramavenkat, Joshua J. Bowers, Santiago A. Lozano-Calderon, David H. Ebb, Kevin A. Raskin, Gregory M. Cote, Edwin Choy, G. Petur Nielsen, Ioannis S. Vlachos, Benjamin Haibe-Kains, Dimitrios Spentzos

## Abstract

Osteosarcoma is a rare primary bone tumor for which no significant therapeutic advancement has been made since the late 1980s despite ongoing efforts. Overall, the five-year survival rate remains about 65%, and is much lower in patients with tumors unresponsive to methotrexate, doxorubicin, and cisplatin therapy. Genetic studies have not revealed actionable drug targets, but our group, and others, have reported that epigenomic biomarkers, including regulatory RNAs, may be useful prognostic tools for osteosarcoma. We tested if microRNA (miRNA) transcriptional patterns mark the transition from a chemotherapy sensitive to resistant tumor phenotype. Small RNA sequencing was performed using 14 patient matched pre-chemotherapy biopsy and post-chemotherapy resection high-grade osteosarcoma frozen tumor samples. Independently, small RNA sequencing was performed using 14 patient matched biopsy and resection samples from untreated tumors. Separately, miRNA specific Illumina DASL arrays were used to assay an independent cohort of 65 pre-chemotherapy biopsy and 26 patient matched post-chemotherapy resection formalin fixed paraffin embedded (FFPE) tumor samples. mRNA specific Illumina DASL arrays were used to profile 37 pre-chemotherapy biopsy and five post-chemotherapy resection FFPE samples, all of which were also used for Illumina DASL miRNA profiling. The National Cancer Institute Therapeutically Applicable Research to Generate Effective Treatments dataset, including PCR based miRNA profiling and RNA-seq data for 86 and 93 pre-chemotherapy tumor samples, respectively, was also used. Paired differential expression testing revealed a profile of 17 miRNAs with significantly different transcriptional levels following chemotherapy. Genes targeted by the miRNAs were differentially expressed following chemotherapy, suggesting the miRNAs may regulate transcriptional networks. Finally, an *in vitro* pharmacogenomic screen using miRNAs and their target transcripts predicted response to a set of candidate small molecule therapeutics which potentially reverse the chemotherapy resistance phenotype and synergize with chemotherapy in otherwise treatment resistant tumors. Importantly, these novel therapeutic targets are distinct from targets identified by a similar pharmacogenomic analysis of previously published prognostic miRNA profiles from pre chemotherapy biopsy specimens.

## INTRODUCTION

Osteosarcoma is the most common primary bone malignancy, with approximately 1,000 new cases per year in the United States (Meltzer & Helman, 2021). Diagnosis is most commonly made in adolescents and young adults, but there is a second incidence peak in later adulthood, the cases of which are often associated with prior radiation exposure. The tumor most often originates in the metaphysis of long bones, with the distal femur and proximal tibia being especially common sites. Location of the tumor in the axial skeleton, such as tumors of the skull or pelvic girdle, is often associated with worse outcome.

Osteosarcoma patients typically first present with pain at the site of the tumor or pathological fracture, for which these patients are especially susceptible because of tumor induced osteolysis (Zhou et al., 2017).

Additionally, we have previously found evidence of more aggressive tumor biology in patients who present with pathologic fractures, who often go on to have worse long-term outcomes (Lozano Calderon et al., 2019). While there are specific imaging findings suggestive of osteosarcoma, definitive diagnosis requires pathological evaluation of a biopsy specimen, often a retrieved through a core needle biopsy (Beird et al., 2022).

Osteosarcomas are typically classified as either low or high grade (Bertoni & Bacchini, 1998). While low grade tumors are generally associated with low metastatic propensity and treated with surgical resection alone, high grade tumors, the focus of the present study, are at an elevated risk of progressing to metastatic disease and are thus additionally treated with chemotherapy.

Known osteosarcoma risk factors are primarily genetic, but specific environmental factors, like prior radiation exposure, also exist (Cole et al., 2022; Mirabello et al., 2009). Cancer predisposition syndromes such as Bloom syndrome, Diamond-Blackfan anemia, Li-Fraumeni syndrome, radial ray defect patellae and palate abnormality diarrhea and dislocated joints limb abnormality little size slender nose and normal intelligence (RAPADILINO) syndrome, retinoblastoma, Rothmund-Thomson Syndrome, and Werner syndrome are known risk factors.

However, only about 28% of patients carry a likely pathogenic germline mutation in a cancer susceptibility gene (Chauveinc et al., 2001; German, 1997; Ishikawa et al., 2000; Larizza et al., 2010; F. P. Li et al., 1988; Lipton et al., 2001; Mirabello et al., 2020; Siitonen et al., 2009). This suggests that other, potentially epigenetic, mechanisms are also important in this disease’s biology. Additionally, most of these mutations are rare, affecting a very small percentage of patients, making efforts to develop therapeutics for these patients more difficult.

For approximately 60-65 percent of high-grade osteosarcoma patients, standard chemotherapeutic treatment and surgical excision of the primary tumor is curative. However, approximately 20 percent of patients present with metastatic disease (Gill & Gorlick, 2021), and in these patients, along with those with unresected or recurrent disease, prognosis is much worse, with long term survival rates at about 30 percent. The current treatment approach for osteosarcoma was developed in the 1980’s, and includes surgical resection of the tumor, and multi-agent chemotherapy including methotrexate, doxorubicin, and cisplatin (MAP) (Rosen et al., 1976, 1979, 1982). Surgical treatment involves complete tumor resection and is often performed with a limb sparing approach rather than amputation. Marginal resections are associated with increased risk of recurrence, but for tumors resected with negative margins, the local recurrence rate is low, at less than five percent (Picci et al., 1994).

Despite ongoing efforts to develop new systemic therapies, little progress in improving disease outcomes has been made since the 1980’s (Beird et al., 2022). The most recent large-scale, international clinical trial in the disease, EURAMOS-1, sought to improve patient outcomes by risk stratifying patients for alternate adjuvant therapies (Whelan et al., 2015). However, the study failed to improve survival in either the high or low risk patient groups (Bielack et al., 2015; Marina et al., 2016). The trial’s failure suggests that the use of pathologically assessed necrosis in the operative specimen following chemotherapy is not a useful tool for risk stratification and drug novel therapeutic application in this disease.

Currently, however, there is a lack of alternative biomarkers for further study. Osteosarcomas are known to have a chaotic genomic landscape, frequently characterized by kaetagis (a large number of localized base pair mutations) and chromothripsis (clustered chromosomal rearrangements), however, few of the genomic aberrations are recurrent or actionable (X. Chen et al., 2014; Lorenz et al., 2016; Perry et al., 2014; Sayles et al., 2019; Yamaguchi et al., 1992). It has been found that some aberrations, such as MYC gene amplification, are prognostic, but these factors have not yet been developed for clinical applications, and generally apply only to a small fraction of patients (Feng et al., 2020; Sayles et al., 2019). Thus, current opinion is that conventional approaches targeting specific single genetic aberrations are unlikely to be successful in osteosarcoma, and other approaches better aligned with biology, potentially including epigenomic markers, are needed (Bishop et al., 2016; Meltzer & Helman, 2021; Roberts et al., 2019).

To this end, we previously reported panels of microRNAs (miRNAs), profiled at the time of diagnostic biopsy, which are prognostic of patient survival (Kelly et al, 2013; Hill et al, 2017; Lietz et al, 2020). MiRNAs are small, about 20 nucleotides long, non-coding RNAs which regulate large fractions of the coded genome post-ranscriptionally. They have been shown to play a variety of roles in cancer (Calin et al., 2005; Calin & Croce, 2006; Esquela-Kerscher & Slack, 2006; J. Lu et al., 2005; Mi et al., 2007; Ooi et al., 2011). While we have previously investigated the static expression of miRNAs in osteosarcoma biopsy specimens, the dynamic changes in these regulatory species induced by chemotherapy have yet to be studied. In the analyses described here, a specific set of miRNAs marking chemotherapy resistance is identified. Pharmacogenomic analysis using the miRNAs provides a set of candidate drugs which may reverse the chemoresistant phenotype, and thus synergize with chemotherapy, providing drug response hypotheses for further study, clinical testing, and future targeted application to improve patient outcomes.

## RESULTS

### The MGH and Longwood miRNA datasets provide for dynamic chemotherapy response testing

Two miRNA transcriptional profiling datasets were generated, both including pre- and post-chemotherapy high grade osteosarcoma specimens from the same patients. The datasets differ with respect to the sample collections dates, type of clinical specimen, and miRNA assay technology (**Table 1**). Herein, we refer to these datasets as the MGH and Longwood paired specimen datasets, with the dataset names reflecting the origin of the patient samples. The MGH and Longwood miRNA datasets contained 7 and 26 pairs of patient-matched pre-chemotherapy biopsy and post-chemotherapy resection samples, respectively. The MGH dataset additionally contained 7 pairs of patient-matched, paired pre-chemotherapy biopsy and pre-chemotherapy resection specimens. Detailed demographic information is presented in **Supplementary Table 1** and summary statistics for the chemoresistant samples, classified as tumors with less than 90 percent necrosis in the operative specimen (Meltzer & Helman, 2021), in **Table 1**. We previously partly reported the samples in the Longwood dataset as part of a separate study testing association between miRNA expression and patient survival using primarily pre-chemotherapy biopsy specimens (Kelly et al., 2013).

**Table 1.**
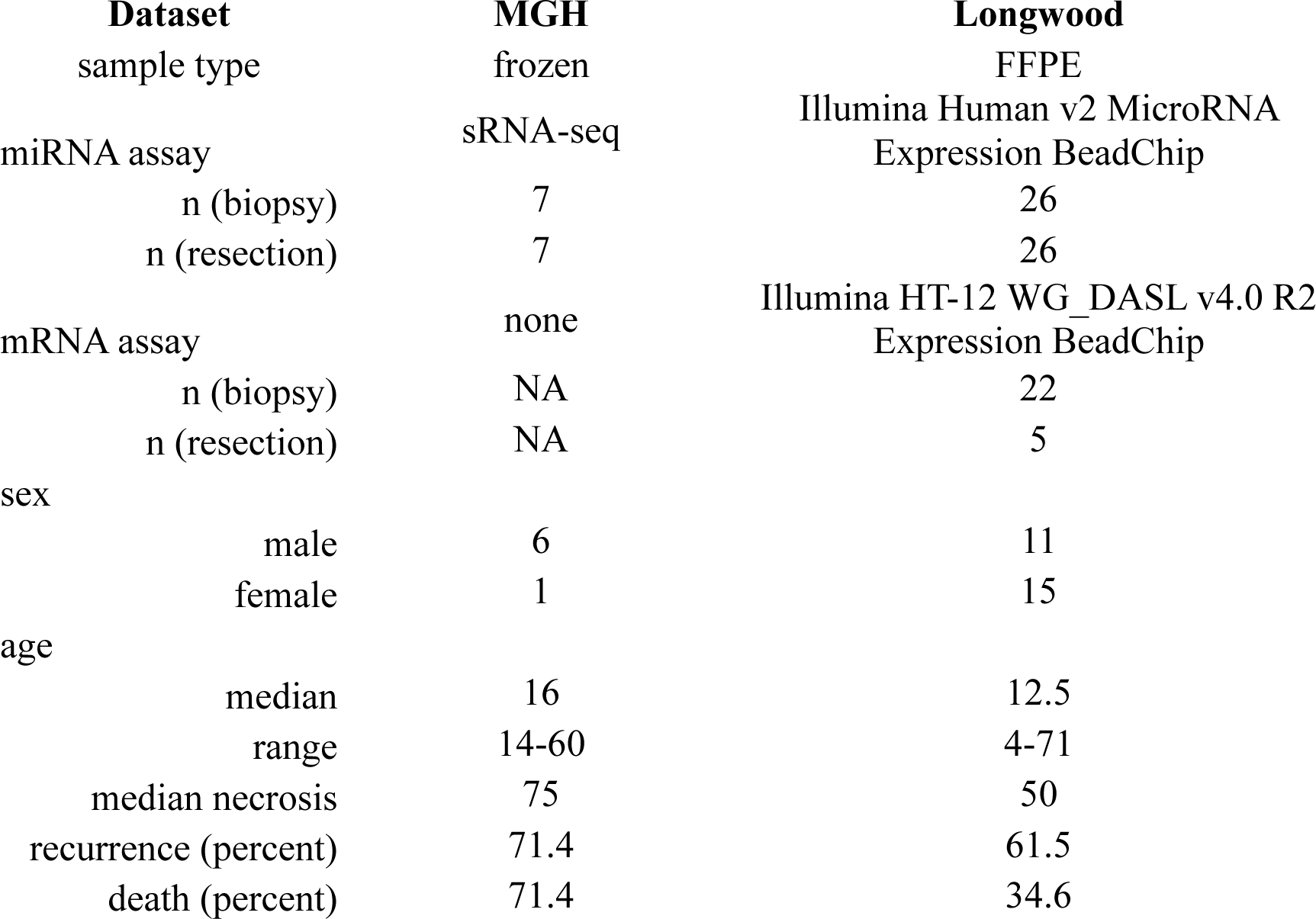
Characteristics of the MGH and Longwood datasets.

In general, both the MGH and Longwood cohorts were composed predominately of adolescent patients, and the cohorts’ age distributions were similar. The MGH cohort contained a larger fraction of male patients.

Chemotherapy regimens were similar in both cohorts, with most patients receiving at least MAP. A small fraction of patients only received Adriamycin (doxorubicin) and cisplatin (AP), and others additionally received adjuvant etoposide and ifosfamide as front-line therapy. The majority of patients in both cohorts underwent the standard treatment course of neoadjuvant therapy, surgical resection, and adjuvant therapy, except in the minority of cases where surgery was performed prior to induction chemotherapy, such as in the case of a pathologic fracture. In the MGH cohort, these unique cases provided both a biopsy and a resection specimen prior to the initiation of chemotherapy, offering the opportunity to control for other biologic or technical differences between biopsy and resection specimens when assessing the effect of chemotherapy on miRNA profile changes over time.

While histological subtype information was not available for the Longwood cohort, in the MGH cohort, five of the seven chemoresistant samples were of the most common osteoblastic subtype, and the remaining two were of the chondroblastic subtype. While histologic subtype is not currently used as a predictive or prognostic tool, it has been suggested that osteoblastic and chondroblastic histologies tend to be more clinically aggressive (Bacci et al., 2003). Both the Longwood and MGH cohorts have relatively long follow up times at 42 and 78 months, respectively, for censored patients.

The MGH dataset was generated using fresh frozen tissue samples, and small RNAs were profiled using small RNA sequencing. The Longwood dataset was generated using formalin fixed paraffin embedded (FFPE) samples, and small RNAs were profiled using Illumina cDNA mediated annealing, selection, extension, and ligation (DASL) arrays, which were optimized for partially degraded RNA, such as that from FFPE samples (Fan et al., 2004).

### Identification of a Consensus Dynamic Chemoresponse miRNA Profile

The paired nature of the MGH and Longwood samples with respect to chemotherapy were first leveraged to test if specific miRNAs may be markers of a chemoresistant phenotype, with the reasoning that the molecular readout from the post-chemotherapy specimens would represent a chemotherapy resistant cell population, as they had survived treatment. Given that the MGH dataset was generated using frozen samples and small RNA sequencing, and the Longwood dataset was generated using FFPE samples and DASL microarray technology, miRNAs with similar patterns of differential expression in both datasets, despite these technical differences, may be robust markers of an OSA chemotherapy resistance phenotype. The 321 miRNAs which passed filtering criteria and could be mapped to both datasets were used for statistical analysis. Note that the DASL microarray platform used in the Longwood dataset interrogates only a subset of miRNAs and miRNA expression is tissue specific, thus the total number of analyzed miRNAs is less than the approximately 2,500 coded in the human genome (Kozomara et al., 2019).

*DESeq2* based paired differential expression analysis was used to test for miRNA transcriptional differences pre- and post-chemotherapy in the MGH sequencing dataset (Love et al., 2014). Analysis was performed, separately, using the 14 chemotherapy treated samples and the 14 untreated samples, which served to control for miRNA transcriptional differences between biopsy and resection specimens not related to chemotherapy. MiRNAs with p < 0.05 were considered differentially expressed. False discovery rate (FDR) corrected p values were not considered in this initial discovery analysis given the small sample size and that the resulting miRNAs will be further tested in independent datasets. 50 miRNAs were differentially expressed following chemotherapy in the MGH dataset (**Fig. 1**), whereas 19 miRNAs were differentially expressed in the untreated group of control pairs with the same sample size. Despite the small sample size, 23 of the 50 differentially expressed miRNAs still had an FDR less than 0.1 (Benjamini & Hochberg, 1995). The larger number of significant findings in the treated compared to untreated pairs served as initial evidence that the discovered molecular differences were due to chemotherapy rather than other factors. Additionally, only two of the miRNAs differentially expressed in the untreated samples were differentially expressed in the treated samples but the direction of change was opposite, further suggesting the observed changes in treated samples may be due to chemotherapy.

**Figure 1.**
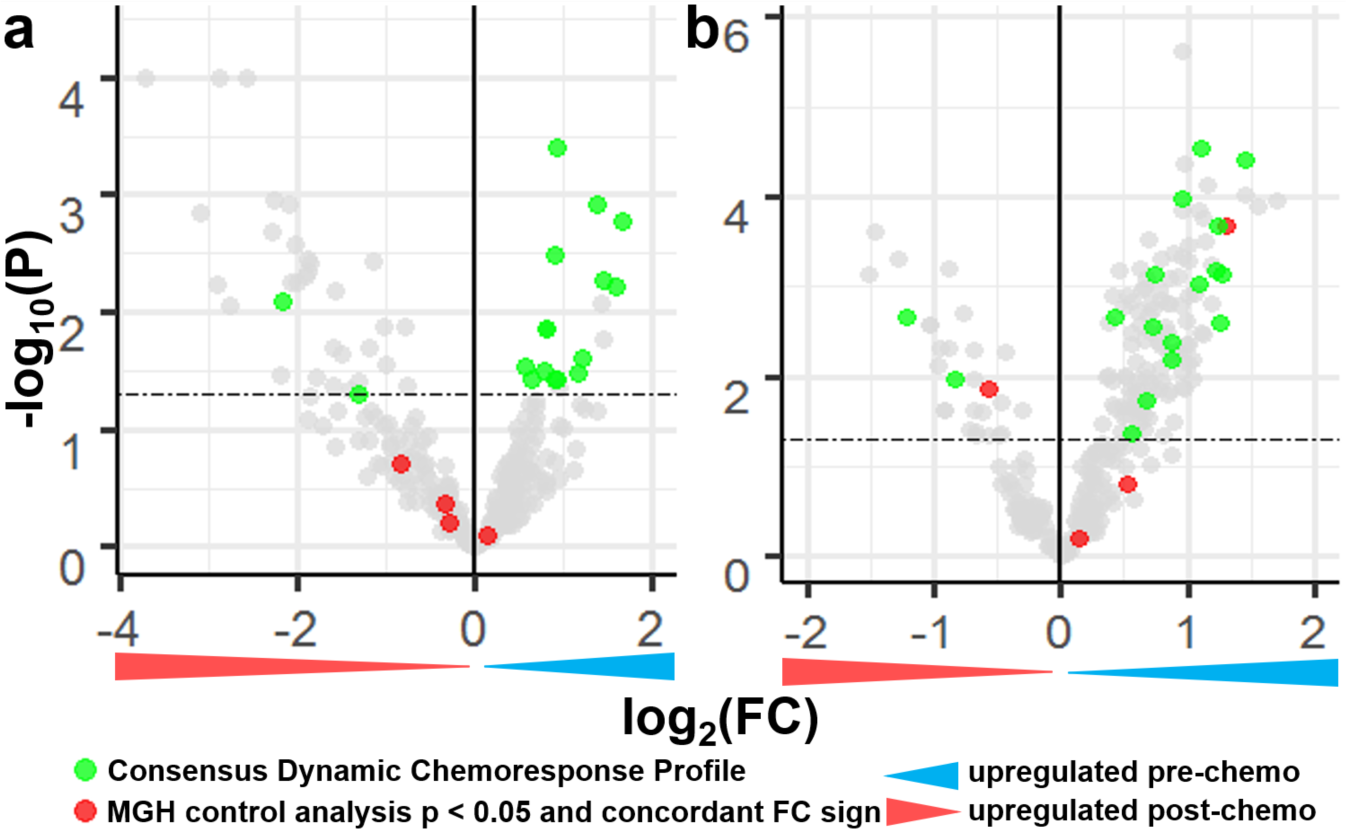
Defining a consensus dynamic chemoresponse profile through miRNA differential expression analysis in the MGH and Longwood Datasets. **a** Volcano plot of paired differential miRNA expression between biopsy and resection samples from chemotherapy treated patients in the MGH dataset. **b** Volcano plot of paired differential miRNA expression between biopsy and resection samples from chemotherapy treated patients in the Longwood dataset. Dashed lines at p = 0.05.

*Limma* based paired differential expression analysis was used to test for miRNA transcriptional differences between pre and post chemotherapy specimens in the Longwood microarray dataset (Ritchie et al., 2015). 166 miRNAs were differentially expressed (**Fig. 1b**), and all 166 also had an FDR corrected p value less than 0.1. A larger number of significant findings were identified in the Longwood dataset potentially because it has about four times as many samples, and thus is more statistically powerful. Only two of the 166 differentially expressed miRNAs were significant and had the same direction of change between the MGH untreated samples.

To identify the most robust miRNA markers of chemotherapy resistance, the intersection of miRNAs differentially expressed following chemotherapy in both datasets was identified. 17 of the 18 identified miRNAs had the same direction of fold change between two datasets (hypergeometric test, p = 6.868 x 10^-6^), suggesting the differentially expressed miRNAs were reproducible genomic changes. These 17 miRNAs were thus considered candidate biomarkers for further testing and are hereafter referred to as the Consensus Dynamic Chemotherapy Response Profile (CDCRP) miRNAs. None of the CDCRP miRNAs were differentially expressed between the untreated MGH samples (**Fig. 1**).

The 17 CDCRP miRNAs were encoded across the genome, without enrichment for a specific locus (**Table 2**). While the majority of the CDCRP miRNAs had greater expression in the pre-chemotherapy samples, two were measured at increased levels in the post-chemotherapy specimens. This increased confidence in the biologic origin of the miRNA differences, as unidirectional changes could have reflected overall decreased miRNA levels in the more necrotic post-chemotherapy resection specimen, as well as artifacts related to surgical resection of the tumor. The two miRNAs with elevated transcriptional levels post-chemotherapy were the 5’ and 3’ arms derived from the same precursor miRNA, mir-654, processed from a primary miRNA encoded on the 14q32 locus. We and others have previously found that a collection of miRNAs from this locus is predictive of patient survival in osteosarcoma (Hill et al., 2017; Kelly et al., 2013; Lietz et al., 2020; Sarver et al., 2013). However, none of the CDCRP miRNAs overlapped with our previously published profile of prognostic miRNAs, suggesting that different biological pathways are involved in chemotherapy response and long-term outcome. There also existed initial evidence that each of the 17 CDCRP miRNAs are connected to chemotherapy response (Bhatia et al., 2019; Cao et al., 2016; Y. Chen et al., 2019; Duan et al., 2017; Hou et al., 2014; Ji et al., 2018; Jin et al., 2019; Lai et al., 2019; H.-Y. Li et al., 2017; M. Lu et al., 2018; Niu et al., 2020; Schreiber et al., 2016; Tie et al., 2018; Wang et al., 2018; M. Xu et al., 2018; Zhang et al., 2018; Zhao et al., 2017; Zhu et al., 2020). This included many connections to MAP based chemotherapy, including reports finding that at least 10 of the miRNAs are associated with cisplatin response, and at least 4 of the miRNAs are associated with doxorubicin response.

**Table 2.** The 17 CDCRP miRNAs. Fold change (FC) color coding is carried over from **Fig. 1**. FC values were calculated as pre-chemotherapy / post-chemotherapy, so positive log_2_(FC) values indicate greater expression in the pre-chemotherapy samples (blue), and negative log_2_(FC) indicate greater expression in the post-chemotherapy samples (red).

| miRNA | locus | $\text{Log}_2(\text{FC}_{\text{MGH}})$ | $\text{Log}_2(\text{FC}_{\text{Longwood}})$ | $\text{p}_{\text{MGH}}$ | $\text{p}_{\text{Longwood}}$ | *in vitro*<br>chemotherapy<br>associations |
| --- | --- | --- | --- | --- | --- | --- |
| miR-103a-3p | 5q34 / 20p13 | 0.9 | 1.2 | 4.E-04 | 2.E-04 | cisplatin |
| miR-454-3p | 17q22 | 1.4 | 1.1 | 0.001 | 3.E-05 | doxorubicin |
| miR-130b-3p | 22q11.21 | 1.7 | 1.5 | 0.002 | 4.E-05 | cisplatin |
| miR-98-5p | Xp11.22 | 0.9 | 1.3 | 0.003 | 0.003 | cisplatin |
| **miR-210-3p** | 11p15.5 | 1.5 | 1.1 | 0.005 | 0.001 | cisplatin |
| miR-105-5p | Xq28 / Xq28 | 1.6 | 0.7 | 0.006 | 0.003 | cisplatin |
| miR-654-3p | **14q32.31** | -2.2 | -1.2 | 0.008 | 0.002 | cisplatin |
| **miR-374b-5p** | Xq13.2 | 0.8 | 1.3 | 0.014 | 0.001 | cisplatin |
| **miR-15b-3p** | 3q25.33 | 0.8 | 0.7 | 0.014 | 0.001 | 5-FU |
| **miR-421** | Xq13.2 | 1.2 | 0.9 | 0.025 | 0.004 | doxorubicin |
| **let-7d-5p** | 9q22.32 | 0.6 | 1.2 | 0.029 | 0.001 | cisplatin |
| **let-7f-5p** | Xp11.2 / 9q22.32 | 0.8 | 0.9 | 0.031 | 0.006 | 5-FU |
| miR-149-5p | 2q37.3 | 1.2 | 0.7 | 0.033 | 0.018 | cisplatin |
| **miR-15b-5p** | 3q25.33 | 0.9 | 1.0 | 0.037 | 1.E-04 | doxorubicin |
| miR-30d-5p | 8q24.22 | 0.6 | 0.4 | 0.037 | 0.002 | 5-FU |
| miR-7-5p | 15q26.1 / 19p13.3 / 9q21.32 | 0.9 | 0.6 | 0.037 | 0.044 | doxorubicin |
| miR-654-5p | **14q32.31** | -1.3 | -0.8 | 0.049 | 0.011 | cisplatin |

The CDCRP was also able to differentiate between pre- and post-chemotherapy specimens in the MGH and Longwood datasets, suggesting that variation between pre and post chemotherapy samples was greater than across patient variation (**Fig. 2**). Hierarchical clustering of the samples using the CDCRP miRNAs generated two groups of samples significantly associated with chemotherapy treatment in both the MGH and Longwood datasets (MGH: X^2^ = 4.667, Odds Ratio (OR) = 5.000, p = 0.031; Longwood: X^2^ = 8.694, OR = 2.174, p = 0.004), suggesting that the CDCRP miRNAs are useful markers of chemotherapy response in combination and that the observed changes induced by chemotherapy are relatively consistent across patients. While the CDCRP miRNAs were defined in these two datasets, the profile was not guaranteed to differentiate between pre- and post-chemotherapy specimens across the entire dataset, as within patient changes were used to generate the profile, and the component of across patient variability could have driven the clustering patterns. While the clustering patterns were imperfect, it was observed that when pre- and post-chemotherapy were grouped together, they were, in some cases, found to be most molecularly similar to the sample from the same patient, indicating that there are patient level differences CDCRP miRNAs. Additionally, the CDCRP miRNAs did not differentiate between the untreated MGH samples (X^2^ = 0.424, OR = 0.833, p = 0.515, **Supplementary Figure 1**).

**Figure 2.**
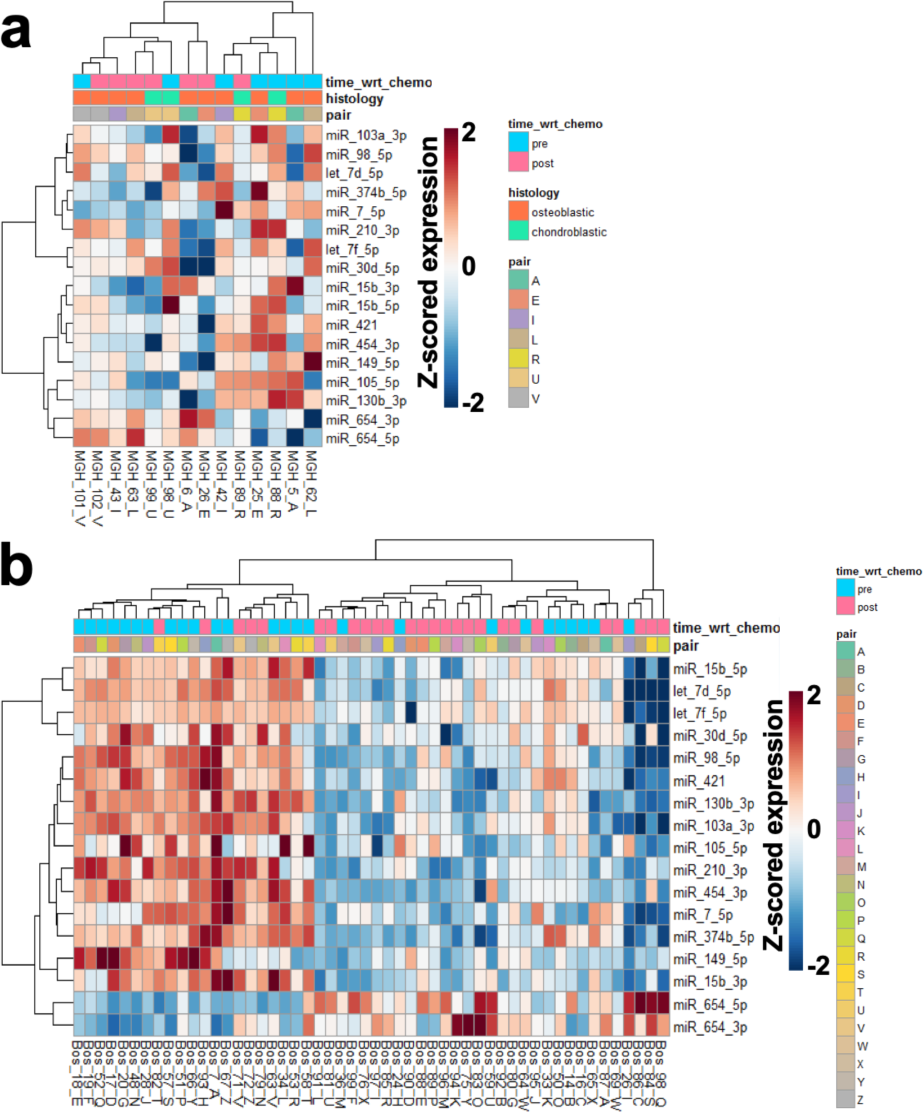
CDCRP expression profiles in the MGH and Longwood datasets. **a** Hierarchical clustering of the MGH chemotherapy treated cohort using expression levels of the CDCRP miRNAs. **b** Hierarchical clustering of the Longwood cohort using expression levels of the CDCRP miRNAs.

### Gene targets of the CDCRP miRNAs have altered transcriptional patterns following chemotherapy

The genes targeted by the CDCRP miRNAs were defined using the DIANA-microT-CDS target prediction algorithm using the DIANA-microTv5.0 server using a microT threshold of 0.7 (Paraskevopoulou et al., 2013). The median number of computationally predicted targets per CDCRP miRNA was 960, and the range was from 16 to 1847 targets. Computationally predicted targets were used because they are not limited by the incompleteness of experimental miRNA target validation, potential biases introduced by performing these experiments in specific tissue types, or targets which may be of current research interest. The CDCRP miRNAs were correlated with their predicted targets to identify a refined list of genes which may be more biologically relevant in osteosarcoma, with the understanding that not all predicted target interactions occur *in vivo*, and that relevant miRNA - gene interactions may be tissue type specific. As the best understood role of miRNAs is to induce mRNA translocation to processing bodies (P-bodies) and subsequent mRNA degradation (Jonas & Izaurralde, 2015; Liu et al., 2005), anti-correlated miRNA and mRNA levels were considered significant (Pearson correlation, one-tailed p < 0.05).

MiRNA – gene target correlations were performed in both the expanded set of biopsy specimens previously reported in the Longwood dataset (Kelly et al., 2013), as well as the NCI sponsored publicly available TARGET osteosarcoma dataset, which provided both TaqMan rt-PCR miRNA data and RNAseq gene expression data for 84 samples. Both datasets were used to identify a set of osteosarcoma relevant set of gene targets with reproducible relationships across different assay platforms and sample types. The TARGET dataset was used in lieu of the MGH dataset, for which only small RNAseq data was generated.

Raw miRNA expression and RNAseq data was downloaded from the TARGET data portal and processed using the 2^-ΔCt^ and *DESeq2* based median of ratios approaches, respectively (Livak & Schmittgen, 2001; Love et al., 2014). 16 of the 17 CDCRP miRNAs were interrogated by the TaqMan platform. The percent of gene targets significantly anticorrelated with each CDCRP miRNA ranged from 2.26 – 15.87% (**Supplementary Table 2**).

In the Longwood dataset, the correlation analysis was performed excluding the subset of samples for which both pre- and post-chemotherapy gene expression data were generated (n = 5), such that this subgroup could later be used as an independent statistical testing environment for candidate genes which we found anticorrelated with their targeting CDCRP miRNA. This provided 32 samples for correlation analysis in the Longwood dataset. The percent of gene targets significantly anticorrelated with each CDCRP miRNA ranged from 2.92 – 14.72% (**Supplementary Table 2**). Individual miRNA-gene target correlations were not correlated (rho < 0.001, p > 0.999) in the MGH and Longwood datasets, highlighting the value of searching for gene targets which display expected patterns of regulation to be further developed as markers rather than using the entire predicted target set.

222 CDCRP miRNA – predicted target pairs were found to be significantly anticorrelated in both the MGH and Longwood datasets, including 198 unique genes and 15 of 16 evaluable CDCRP miRNAs (one of the CDCRP miRNAs was not interrogated by the TaqMan platform used to generate the TARGET dataset). Limma based differential expression testing was then performed using this set of genes in the five pairs of pre- and post-chemotherapy samples in the Longwood dataset for which gene expression data was generated. None of these pairs, neither pre- nor post-chemotherapy sample, was used for correlation analysis, and they thus represent an independent test of chemotherapy induced gene expression changes. 30 of the anticorrelated miRNAs were differentially expressed, and 28 of 30 were differentially expressed with the expected log fold change sign based on dynamic change in miRNA expression, with the expected sign being opposite of that of the targeting miRNA, suggesting that the CDCRP miRNAs may be involved in a larger regulatory network relevant to chemotherapy response (hypergeometric test, p = 1.4 x 10^-8^; **Fig. 3**; **Table 3**).

**Figure 3.**
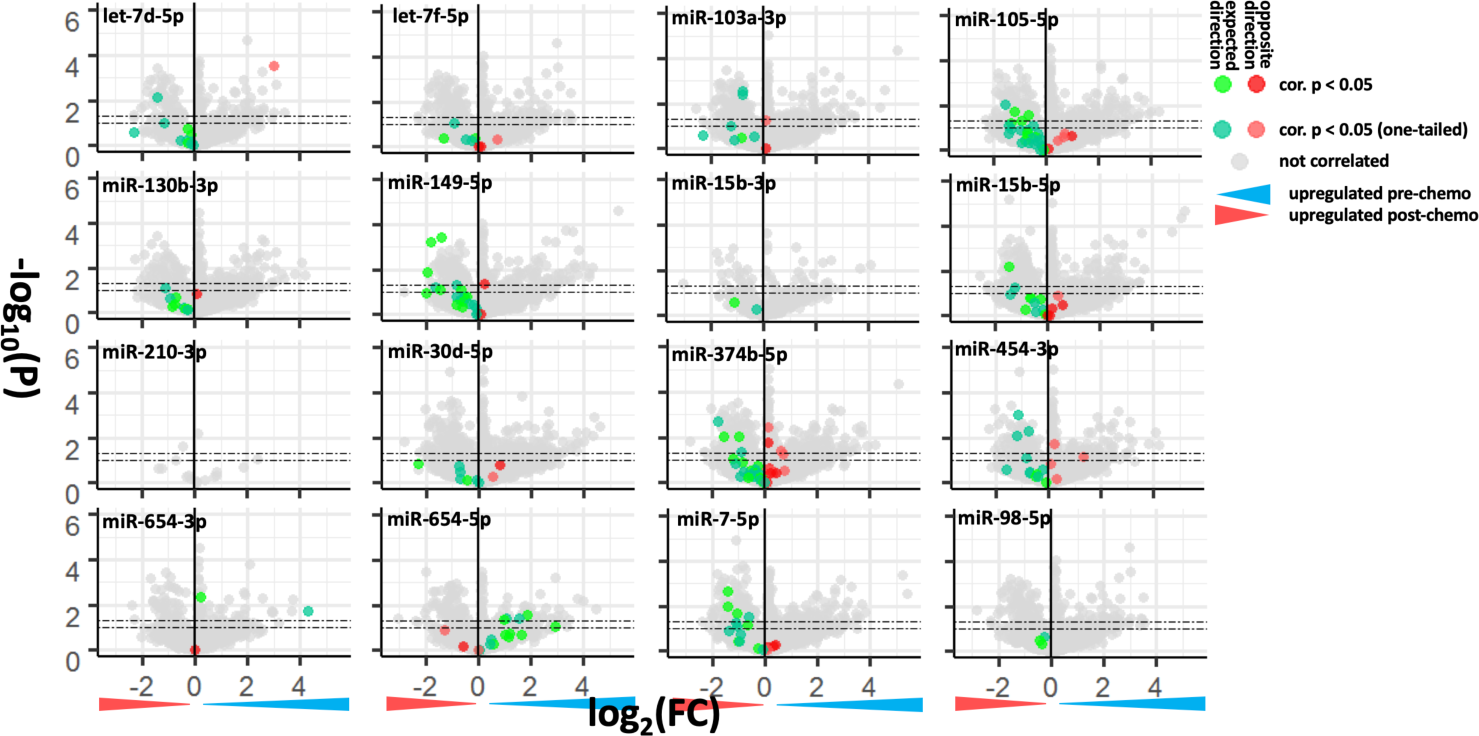
Differential expression of the CDCRP miRNA anticorrelated gene targets in the five chemotherapy treated pairs with both miRNA and gene expression profiling in the Longwood dataset which were not used for correlation analysis.

**Table 3.** CDCRP miRNA anticorrelated gene targets significantly differentially expressed (DE) with expected fold change (FC) direction between pre and post chemotherapy specimens in the Longwood (LW) dataset.

| miR | gene | DE log2(FC) | DE p | TARGET r | TARGET p | LW r | LW p |
| --- | --- | --- | --- | --- | --- | --- | --- |
| miR_15b_5p | SLC29A2 | -1.4739 | 0.0065 | -0.2537 | 0.0099 | -0.4121 | 0.0096 |
| miR_15b_5p | DNAJC5 | -1.2176 | 0.0550 | -0.2124 | 0.0262 | -0.2998 | 0.0478 |
| miR_130b_3p | FASTK | -1.1203 | 0.0741 | -0.1900 | 0.0418 | -0.3724 | 0.0179 |
| miR_105_5p | BRIP1 | -1.5564 | 0.0089 | -0.1901 | 0.0417 | -0.4909 | 0.0022 |
| miR_105_5p | WAC | -1.1844 | 0.0192 | -0.2615 | 0.0081 | -0.4284 | 0.0072 |
| miR_105_5p | NAA50 | -1.3219 | 0.0635 | -0.2208 | 0.0218 | -0.4156 | 0.0090 |
| miR_105_5p | PPIE | -1.4344 | 0.0736 | -0.2016 | 0.0329 | -0.3372 | 0.0296 |
| let_7d_5p | ZFYVE16 | -1.3959 | 0.0076 | -0.1816 | 0.0491 | -0.3560 | 0.0228 |
| miR_654_5p | TANK | 1.8639 | 0.0261 | -0.3266 | 0.0012 | -0.4712 | 0.0032 |
| miR_654_5p | EFNA1 | 1.5361 | 0.0390 | -0.4377 | 1.57E-05 | -0.3133 | 0.0404 |
| miR_654_5p | PTPRD | 1.0426 | 0.0405 | -0.2364 | 0.0152 | -0.3230 | 0.0357 |
| miR_654_5p | TNK2 | 2.9069 | 0.0878 | -0.2890 | 0.0038 | -0.3612 | 0.0211 |
| miR_149_5p | LARP4 | -1.4344 | 0.0004 | -0.2573 | 0.0091 | -0.4783 | 0.0028 |
| miR_149_5p | PANX1 | -1.8365 | 0.0006 | -0.2624 | 0.0079 | -0.4777 | 0.0028 |
| miR_149_5p | FLNA | -1.9434 | 0.0134 | -0.2728 | 0.0060 | -0.6079 | 0.0001 |
| miR_149_5p | CHCHD3 | -1.6439 | 0.0631 | -0.2186 | 0.0229 | -0.3452 | 0.0265 |
| miR_149_5p | PHACTR4 | -1.4739 | 0.0823 | -0.2207 | 0.0218 | -0.6056 | 0.0001 |
| miR_454_3p | WDR47 | -1.1520 | 0.0010 | -0.2070 | 0.0294 | -0.5642 | 0.0004 |
| miR_454_3p | RTKN | -1.1844 | 0.0079 | -0.2139 | 0.0254 | -0.4173 | 0.0087 |
| miR_7_5p | CAMKK2 | -1.3959 | 0.0022 | -0.2738 | 0.0059 | -0.6287 | 0.0001 |
| miR_7_5p | ZMAT3 | -1.4344 | 0.0098 | -0.2196 | 0.0224 | -0.5634 | 0.0004 |
| miR_7_5p | WDFY3 | -1.0589 | 0.0202 | -0.2470 | 0.0117 | -0.4016 | 0.0114 |
| miR_7_5p | POLH | -1.1203 | 0.0552 | -0.1820 | 0.0488 | -0.6536 | 2.49E-05 |
| miR_7_5p | TNRC6B | -1.0589 | 0.0752 | -0.2007 | 0.0336 | -0.3168 | 0.0386 |
| miR_374b_5p | ZNF644 | -1.7859 | 0.0020 | -0.2039 | 0.0314 | -0.3173 | 0.0384 |
| miR_374b_5p | BRIP1 | -1.5564 | 0.0089 | -0.2773 | 0.0053 | -0.4662 | 0.0036 |
| miR_374b_5p | LYSMD3 | -1.1844 | 0.0875 | -0.3364 | 0.0009 | -0.5243 | 0.0010 |
| miR_654_3p | HEY2 | 4.2965 | 0.0190 | -0.3309 | 0.0011 | -0.3422 | 0.0276 |

### Gene targets of the CDCRP miRNAs provide additional insight into the biology of chemotherapy resistance

Functional annotation analysis of the genes predicted to be targets of the CDCRP miRNAs was performed using DIANA-miRPath v3.0 (Vlachos et al., 2015). Gene sets from the Kyoto Encyclopedia of Genes and Genomes (KEGG) and Gene Ontology (GO) databases were analyzed on the DIANA web server using both standard hypergeometric distributions and more stringent unbiased empirical distributions to account for biases encountered by traditional enrichment statistics with miRNA target gene sets (Ashburner et al., 2000; Bleazard et al., 2015; Consortium, 2021; Kanehisa et al., 2019, 2023; Kanehisa & Goto, 2000). Meta analysis statistics, which provides a single pooled enrichment statistic for pathways enriched in target sets of multiple miRNAs, are also employed to account for the combined effect of multiple miRNAs targeting the same set of pathways. 31 KEGG pathways and 69 GO categories were found significant (FDR < 0.05) by the hypergeometric test. Five and zero of the KEGG and GO terms were also significant in the empirical distribution tests, respectively, potentially representing regulated pathways most specific to the CDCRP miRNAs (**Fig. 4**).

**Figure 4.**
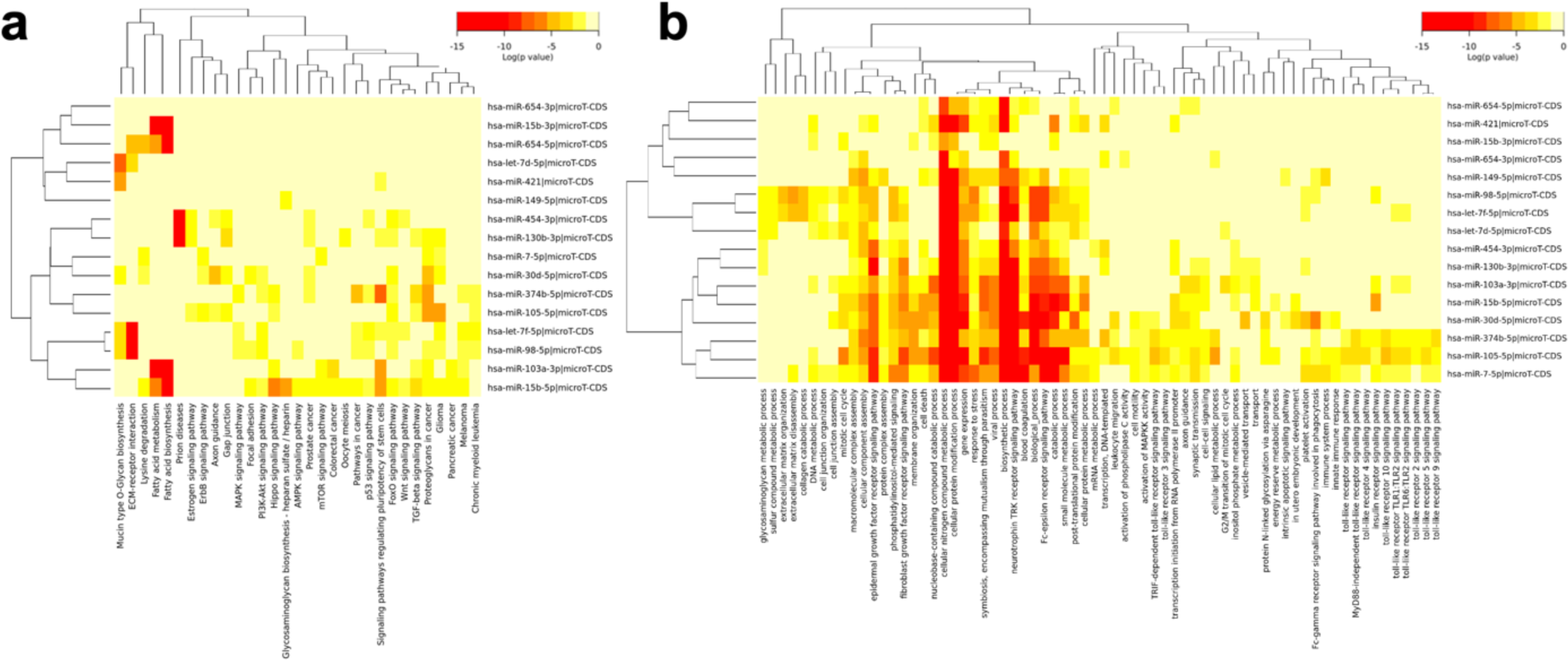
Clustered heatmap of **a** KEGG and **b** GO terms significantly enriched (FDR < 0.05) in the CDCRP miRNA gene target set by miRPath hypergeometric meta-analysis.

Pathways potentially related to chemoresistance, such as the KEGG pathway “Signaling Pathways Regulating Pluripotency of Stem Cells”, were identified, invoking the idea that the chemoresistant cell population may harbor cancer stem cells (Batlle & Clevers, 2017), which have been shown to be responsible for treatment resistance in many tumor types, including osteosarcoma (Brown et al., 2017).

### The CDCRP miRNAs and genes predict response to novel therapeutics

Given that the CDCRP miRNAs were found to mark the emergence of chemoresistance to standard methotrexate, doxorubicin, and cisplatin-based therapy (MAP), it was tested if they could also mark the sensitivity to novel therapeutics which may be useful in resistant tumors. These therapeutics may then be used to synergize with MAP. The large-scale Profiling Relative Inhibition Simultaneously in Mixtures (PRISM) drug screening dataset, which profiled the sensitivity of 930 genomically characterized cell lines to 4,518 drugs, was first leveraged to test if static levels of the CDCRP miRNAs predict response to novel agents (Corsello et al., 2020; Yu et al., 2016). Drug dose-response sensitivity from the PRISM secondary screen and miRNA data from the Cancer Cell Line Encyclopedia (CCLE) project, which genomically characterized the same cell lines used in the PRISM drug response screen, were downloaded using the Cancer Dependency Map (DepMap) data portal (Barretina et al., 2012; Ghandi et al., 2019; Tsherniak et al., 2017). Cell lines were randomly assigned to independent training sets (2/3 of cell lines) and test sets (1/3 of cell lines).

Simple linear regression using the CDCRP miRNAs to predict AUC values was performed using the *glmnet* R package (Friedman et al., 2010; Tibshirani, 1996). Models were trained for drugs with at least 20 cell lines in the training set. 31 miRNA drug combinations were found to have significant predictive capacity in the training set (FDR < 0.05), significant and positive correlation between prediction and actual AUC in the testing set (p < 0.05), and a positive correlation between prediction and actual AUC in the small subset of OSA cell lines (**Table 4**). Definitive statistical analysis could not be performed for the very few osteosarcoma cell lines, at most five, and the osteosarcoma cell lines were intentionally not excluded from the training set such that these most informative datapoints could be used to train the models.

**Table 4.** CDCRP miRNA based predictive drug response model performance in the training, test, and osteosarcoma (OSA) cell line sets.

| miRNA | drug | MoA | target | coef | training FDR | test cor | test p | OSA cor | n (-)<br>connectivity<br>hits |
| --- | --- | --- | --- | --- | --- | --- | --- | --- | --- |
| 30d-5p | vinblastine | tubulin polymerization inhibitor | JUN, tubulin | 0.327 | 5.64E-08 | 0.258 | 0.002 | 0.575 | 9 |
| 30d-5p | verubulin | tubulin polymerization inhibitor | TUBB | 0.287 | 6.35E-06 | 0.256 | 0.003 | 0.619 | NA |
| 30d-5p | BNC105 | tubulin polymerization inhibitor | NA | 0.289 | 1.52E-05 | 0.209 | 0.015 | 0.473 | NA |
| 30d-5p | VU0238429 | acetylcholine receptor allosteric modulator | CHRM5 | 0.286 | 2.74E-05 | 0.180 | 0.041 | 0.401 | NA |
| 30d-5p | vindesine | tubulin polymerization inhibitor | TUBB, TUBB1 | 0.259 | 1.88E-04 | 0.275 | 0.001 | 0.483 | 2 |
| let-7d-5p | NSC-697923 | ubiquitin-conjugating enzyme inhibitor | UBE2N | 0.195 | 0.001 | 0.297 | 4.56E-04 | 0.274 | 0 |
| let-7f-5p | ryuvidine | histone lysine methyltransferase inhibitor, CDK inhibitor | KDM5A, CDK2, CDK4 | 0.167 | 0.001 | 0.283 | 0.001 | 0.696 | 7 |
| 654-3p | canertinib | EGFR inhibitor | AKT1, EGFR, ERBB2, ERBB4 | -0.784 | 0.001 | 0.381 | 1.19E-04 | 0.908 | 10 |
| let-7f-5p | NSC-697923 | ubiquitin-conjugating enzyme inhibitor | UBE2N | 0.165 | 0.001 | 0.179 | 0.037 | 0.411 | 0 |
| 30d-5p | axitinib | PDGFR nhibitor, VEGFR inhibitor | CSF1, FLT1, FLT4, KDR, PLK4 | 0.286 | 0.001 | 0.290 | 0.005 | 0.127 | 2 |
| 30d-5p | fluvastatin | HMGCR inhibitor | HMGCR | 0.235 | 0.001 | 0.302 | 3.53E-04 | 0.513 | 2 |
| 654-3p | lapatinib | EGFR inhibitor | EGFR, ERBB2 | -0.799 | 0.001 | 0.271 | 0.013 | NA | 19 |
| 374b-5p | fluvastatin | HMGCR inhibitor | HMGCR | 0.228 | 0.004 | 0.242 | 0.005 | 0.124 | 2 |
| let-7d-5p | ryuvidine | histone lysine methyltransferase inhibitor, CDK inhibitor | CDK2, CDK4 | 0.173 | 0.005 | 0.238 | 0.006 | 0.651 | 7 |
| 103a-3p | CGS-15943 | adenosine receptor antagonist | ADORA | -0.273 | 0.005 | 0.373 | 2.66E-04 | NA | 1 |
| 103a-3p | fluvastatin | HMGCR inhibitor | HMGCR | 0.212 | 0.006 | 0.332 | 8.06E-05 | 0.551 | 2 |
| 30d-5p | selinexor | exportin antagonist | XPO1 | 0.211 | 0.006 | 0.284 | 0.001 | 0.754 | NA |
| 30d-5p | daunorubicin | topoisomerase inhibitor | TOP2A, TOP2B | 0.224 | 0.006 | 0.179 | 0.048 | 0.756 | 17 |
| 30d-5p | vinorelbine | tubulin polymerization inhibitor | tublin | 0.210 | 0.009 | 0.293 | 0.001 | 0.008 | 8 |
| 130b-3p | Ro-90-7501 | NA | APP | -0.553 | 0.009 | 0.603 | 0.002 | NA | 3 |
| 421 | CGS-15943 | adenosine receptor antagonist | ADORA | -0.418 | 0.013 | 0.225 | 0.032 | NA | 1 |
| 454-3p | RAF265 | RAF inhibitor, VEGFR inhibitor | BRAF | 0.311 | 0.013 | 0.326 | 4.08E-04 | NA | NA |
| let-7d-5p | gambogic-acid | caspase activator | BCL2 | 0.165 | 0.015 | 0.177 | 0.043 | 0.757 | NA |
| 454-3p | hesperadin | Aurora kinase inhibitor | AURKB | 0.318 | 0.017 | 0.212 | 0.031 | NA | NA |
| 654-3p | ibrutinib | Bruton's tyrosine kinase (BTK) inhibitor | BLK, BMX, BTK | -0.689 | 0.027 | 0.323 | 0.006 | NA | 11 |
| 30d-5p | R547 | CDK inhibitor | CDK1, CDK2, CDK4, CDK7 | 0.206 | 0.029 | 0.316 | 0.001 | 0.225 | NA |
| 210-3p | erastin | ion channel antagonist | VDAC2 | -0.198 | 0.029 | 0.322 | 0.001 | 0.957 | 3 |
| 421 | givinostat | HDAC inhibitor | pan-HDAC | -0.337 | 0.032 | 0.208 | 0.017 | 0.563 | 5 |
| 454-3p | BNC105 | tubulin polymerization inhibitor | NA | 0.274 | 0.032 | 0.252 | 0.003 | 0.186 | 0 |
| 454-3p | daunorubicin | topoisomerase inhibitor | TOP2A, TOP2B | 0.281 | 0.034 | 0.258 | 0.004 | 0.578 | 17 |
| 98-5p | NSC-697923 | ubiquitin-conjugating enzyme inhibitor | UBE2N | 0.142 | 0.050 | 0.210 | 0.014 | 0.555 | 0 |

While a diverse set of drug mechanisms of action were represented in **Table 4**, multiple agents interfering with microtubule polymerization and stability, aurora kinase inhibitors, and epigenetic modifying agents, such as histone deacetylase and methyltransferase inhibitors were identified. These drug classes may be entry points into future osteosarcoma drug development efforts.

Given that expression levels of the CDCRP miRNAs reflect a change towards a chemoresistant phenotype, drugs which reverse these genomic changes back towards chemosensitivity could be valuable in synergizing with standard chemotherapy. Connectivity Map (CMap) analysis was performed using the CLUE.io web server to test for such drugs in a dynamic setting (Lamb et al., 2006; Subramanian et al., 2005, 2017). The expression datasets used in the pipeline do not include miRNAs, so we used the 27 targets of the CDCRP miRNAs which were found to have the strongest evidence of involvement in the miRNA regulated chemoresistance network as input for the CMap analysis. Of the 23 drugs presented in **Table 4**, perturbation data was available for 15. Fourteen of the 15 drugs had preliminary evidence of a connectivity score that would reverse the chemoresistant phenotype towards the chemosensitive phenotype, given their negative connectivity scores. These proof of principle results suggest the CDCRP miRNAs and their regulatory networks may be actionably targeted by novel therapeutics to reverse MAP based chemotherapy resistance.

### Pre chemotherapy survival associated miRNAs predict response to distinct therapeutics compared to the CDCRP miRNAs

Our previously published prognostic 22-miRNA profile miRNAs were also tested via linear regression using the same CCLE miRNA and PRISM drug response data (Kelly et al, 2013). 19 miRNA drug combinations were found to have significant predictive capacity in the training set (FDR < 0.05), significant and positive correlation between prediction and actual AUC in the testing set (p < 0.05), and a positive correlation between prediction and actual AUC in the small subset of OSA cell lines (**Table 5**). Of the 19 drugs, seven inhibited MEK, including MEK inhibitor PD-0325901 (mirdametinib) which was previously identified in separate gene expression, and methylation-based screens for drugs with effect in OSA (Lietz et al, 2019; Lietz et al, 2022). Additionally, four other drugs acted upstream of MEK in the MAPK pathway through RAF.

**Table 5.** Prognostic 22-miRNA profile based predictive drug response model performance in the training, test, and osteosarcoma (OSA) cell line sets.

| miRNA | drug | MoA | target | coef | training FDR | test cor | test p | OSA cor | n (-)<br>connectivity<br>hits |
| --- | --- | --- | --- | --- | --- | --- | --- | --- | --- |
| 514a-3p | dabrafenib | RAF inhibitor | BRAF, LIMK1, NEK11, RAF1, SIK1 | -0.511 | 2.25E-08 | 0.543 | 6.29E-06 | NA | 4 |
| 514a-3p | vemurafenib | RAF inhibitor | BRAF, RAF1 | -0.500 | 3.48E-08 | 0.405 | 3.41E-04 | NA | 3 |
| 514a-3p | cobimetinib | MEK inhibitor | NA | -0.441 | 1.56E-06 | 0.418 | 8.31E-05 | NA | 3 |
| 514a-3p | binimetinib | MEK inhibitor | MAP2K1, MAP2K2 | -0.410 | 2.53E-06 | 0.389 | 6.91E-06 | 0.353 | NA |
| 514a-3p | TAK-733 | MEK inhibitor | MAP2K1 | -0.397 | 4.13E-06 | 0.254 | 0.004 | 0.766 | 5 |
| 514a-3p | Ro-4987655 | MEK inhibitor | MAP2K1 | -0.397 | 4.13E-06 | 0.349 | 5.83E-05 | 0.214 | 1 |
| 514a-3p | PD-0325901 | MEK inhibitor | MAP2K1 | -0.352 | 2.25E-04 | 0.221 | 0.015 | 0.445 | 11 |
| 125b-5p | CGS-15943 | adenosine receptor antagonist | ADORA1, ADORA2A, ADORA2B, ADORA3 | 0.135 | 3.99E-04 | 0.301 | 0.004 | NA | 3 |
| 125b-5p | tetrandrine | calcium channel blocker | SLC6A3 | 0.105 | 0.002 | 0.255 | 0.003 | 0.468 | 0 |
| 125b-5p | LY2874455 | FGFR antagonist | FGFR1, FGFR2, FGFR3, FGFR4, KDR | -0.103 | 0.002 | 0.254 | 0.003 | 0.230 | NA |
| 514a-3p | CNX-774 | Bruton's tyrosine kinase (BTK) inhibitor | BTK | -0.330 | 0.003 | 0.352 | 4.70E-04 | NA | 0 |
| 514a-3p | RAF265 | RAF inhibitor, VEGFR inhibitor | BRAF | -0.326 | 0.004 | 0.356 | 1.03E-04 | NA | NA |
| 125b-5p | fluvastatin | HMGCR inhibitor | HMGCR | -0.097 | 0.005 | 0.178 | 0.038 | 0.053 | 0 |
| 125b-5p | neratinib | EGFR inhibitor | EGFR, ERBB2, KDR | 0.108 | 0.009 | 0.345 | 3.63E-04 | 0.165 | 8 |
| 514a-3p | AS-703026 | MEK inhibitor | MAP2K1, MAP2K2 | -0.315 | 0.010 | 0.399 | 1.18E-04 | NA | 3 |
| 125b-5p | tucatinib | EGFR inhibitor | ERBB2 | 0.099 | 0.025 | 0.326 | 0.001 | 0.656 | NA |
| 514a-3p | ABT-702 | adenosine kinase inhibitor | ADK | -0.305 | 0.031 | 0.388 | 1.61E-04 | NA | 3 |
| 514a-3p | PD-318088 | MEK inhibitor | MAP2K1 | -0.273 | 0.039 | 0.345 | 3.59E-04 | 0.671 | NA |
| 514a-3p | GDC-0879 | RAF inhibitor | BRAF | -0.337 | 0.048 | 0.623 | 0.001 | NA | 10 |

Of the 19 drugs presented in **Table 5**, perturbation data in CMap was available for 17, and 14 of these drugs had negative connectivity scores, offering preliminary evidence they may reverse a poor prognosis phenotype towards a phenotype with longer survival.

Of the 39 unique drugs in **Table 4** and **Table 5**, only three were identified in both analyses, CGS-15943, RAF265, and fluvastatin. Further examination of the drugs identified in **Table 4** and **Table 5** also reveals that they predominantly act through distinct mechanisms, an observation with significant biologic and therapeutic implications discussed below.

## DISCUSSION

Current treatments for osteosarcoma were developed about four decades ago, and, despite ongoing efforts, patient outcomes have improved little since that time (Meltzer & Helman, 2021). While about 60-65 percent of patients are cured by surgical resection and systemic chemotherapy, the other 40 percent are still subjected to the toxic effects of treatment but see little or no survival benefit as the tumor spreads and becomes the cause of significant morbidity and mortality. There thus exists an unmet need to develop new drugs for patients who do not respond to current therapies.

Regulatory miRNAs were tested as biomarkers for chemotherapy resistance, and candidate markers were developed as tool to stratify patients for additional therapies which may synergize with the MAP regimen. A profile of 17 miRNAs marking the transition to a chemoresistant state, the consensus dynamic chemoresponse signature (CDCRP), was discovered. The profile was contextualized using the pathways potentially regulated by the miRNAs, and it was used to identify a set of novel drugs for future testing. Thus, the CDCRP provides both an insight into chemoresistance and is a possible biomarker to be further developed for application in conjunction with new therapies. While this is the first report describing an in depth genome wide evaluation of the dynamic changes of miRNA expression with chemoresistance in human samples, all 17 of the miRNAs in the CDCRP have individually and independently been reported to be associated with chemotherapy response, and most to at least one of the MAP agents (Bhatia et al., 2019; Cao et al., 2016; Y. Chen et al., 2019; Duan et al., 2017; Hou et al., 2014; Ji et al., 2018; Jin et al., 2019; Lai et al., 2019; H.-Y. Li et al., 2017; M. Lu et al., 2018; Niu et al., 2020; Schreiber et al., 2016; Tie et al., 2018; Wang et al., 2018; M. Xu et al., 2018; Zhang et al., 2018; Zhao et al., 2017; Zhu et al., 2020). This includes the miRNA hsa-miR-15b-5p, which was previously independently studied by our group in the context of osteosarcoma doxorubicin resistance (Duan et al., 2017). In the study, hsa-miR-15b-5p downregulation was also found to be associated with chemoresistance. The relationship was discovered using different methodology, by performing differential expression testing between parent and doxorubicin resistant cell lines, and then the association was validated in a small human cohort. These congruent results support the use of cell lines for further study of our markers, and specifically, their use in the present pharmacogenomic analysis to discover new drugs which may synergize with MAP chemotherapy.

Given the dynamic nature of the CDCRP, it was reasoned that drugs which prevent the transition to the chemoresistant profile may be useful in combination with MAP to enhance response in otherwise resistant tumors. To identify these drug candidates, a pharmacogenomic screen was performed using the CCLE and PRISM miRNA profiling and drug response datasets (Corsello et al., 2020; Ghandi et al., 2019), and identified a set of 13 candidate drugs. A limitation of the study is that these drugs were discovered by associating the static levels of the CDCRP miRNAs in untreated cell lines. Ideally, a perturbational dataset with miRNA profiling before and after treatment would have been used for our predictive modeling analysis. However, no such dataset currently exists. Instead, the L1000 Touchstone CMAP LINCS 2020 dataset (Lamb et al., 2006; Subramanian et al., 2005, 2017), which provides dynamic gene expression data before and after cell line treatment with a broad range or perturbagens, was leveraged. For 90% of the proposed drugs which were tested in the L1000 Touchstone CMAP LINCS 2020 dataset, there was initial evidence that a cell’s gene expression profile can be changed in the opposite direction as that seen during the evolution to a chemoresistant phenotype. This suggested these drugs may synergize with MAP based chemotherapy and potentially reverse or mitigate resistance to chemotherapy.

The candidate drug list largely contained drugs currently being developed as anti-cancer therapies, specifically tubulin inhibitors, kinase inhibitors, and epigenetic modifying agents, despite the fact the PRISM dataset was designed to also include a large fraction of so-called non-oncology drugs (Corsello et al., 2020). Multi-kinase inhibitors and epigenetic modifying agents were recently proposed as top drug classes for further testing in osteosarcoma based on expert review of current experimental evidence (Whittle et al., 2021). Given the alignment of the drug classes coming out of the analysis with those independently hypothesized to have effect in the disease, further testing of the proposed candidate drugs in conjunction with the CDCRP markers may be warranted and could lead to improved tailored application of novel drugs and optimal individual patient outcomes.

Finally, it should be noted that drugs identified in **Table 4** and **Table 5** predominantly act through distinct mechanisms, further suggesting that survival and chemotherapy resistance networks are different, a concept also supported by the non-overlap between the CDCRP miRNAs presented in this study and the miRNAs prognostic of survival in pre-treatment tumors, that we presented in our previous work (Kelly et al, 2013; Lietz et al 2020). Additionally, this observation also suggests that different drugs may be optimal candidates to therapeutically target these different poor outcome phenotypes. Ultimately, promising therapeutic hypotheses identified by the bioinformatic analysis of public experimental *in vitro* data presented here, should be studied in a targeted manner using *in vivo* models, in order to select the best candidates for clinical testing. Our comprehensive bioinformatic analysis presented here, thus substantially facilitates, and expedites this process by providing rational biomarker-based selection and prioritization of drugs for further development.

## METHODS

### Generation of the Massachusetts General Hospital microRNA dataset

All experiments were approved by the Massachusetts General Hospital (MGH). The MGH IRB waived the requirement of informed consent for this retrospective tissue and clinical information protocol which obtained tissue discarded from routine medical care. All experiments were performed in accordance with relevant guidelines and regulations. Twenty-one pairs of samples, one biopsy and one resection specimen from the same patient per pair, frozen tumor samples from patients with high grade osteosarcoma were retrieved from the archives of the Department of Pathology at MGH. Samples were banked between 1998 to 2010. Samples selection was performed chronologically. Hematoxylin and eosin stained slides generated from the frozen high grade osteosarcomas tissue specimens were reviewed by an expert study pathologist to confirm the diagnosis of high grade osteosarcoma and to determine tumor cellularity. Samples for which there was no available tissue or that failed pathology confirmation of high grade osteosarcoma were excluded from the study, leaving 14 pairs for further analysis. Seven pairs of pre-chemotherapy biopsy specimens and post-chemotherapy resection specimens from neoadjuvant chemotherapy treated patients were included for chemotherapy response analysis. Seven pairs of biopsy and resection specimens from patients who did not receive neoadjuvant therapy were used as untreated control pairs.

RNA was isolated with the QIAGEN RNeasy® Plus Universal Mini Kit (Qiagen) using an adapted protocol designed to increase the quality of extracted RNA from frozen bone specimens (Carter et al., 2012). Sequencing was performed at the Center for Cancer Computational Biology at Dana Farber Cancer Institute (Boston, MA). Qubit was used to determine RNA quantity using the Qubit RNA High Sensitivity Assay Kit reagents (Life Tech). Then, RNA quality was assessed using a Bioanalyzer and Agilent RNA Pico Kit. The NEBNext Multiplex Small RNA Library Prep Kit for Illumina (NEB) was used to convert 100 ng of total RNA into a DNA library following the manufacturer’s protocol, without modifications. Qubit High Sensitivity DNA Kit (Life Tech) was used to determine library quantity and Bioanalyzer High Sensitivity Chip Kit (Agilent) was used to determine library size. Additionally, libraries were assessed via qPCR using the Universal Library Quantification Kit for Illumina (Kapa Biosystems) and 7900HT Fast qPCR machine (ABI). All libraries passing quality control were then diluted to 2 nM, combined into library pools, and sequenced on the NextSeq 500 (Illumina) at a final concentration of 2 pM. Sequencing was performed using three NextSeq Single Read 75 Cycle High Throughput V2 flowcells following standard protocols. The sRNAtoolbox sRNAbench tool was then used to process and quantify small RNA reads (Rueda et al., 2015). SRNAtoolbox identified and removed adapter sequences from the input fastq files. Hierarchal read mapping was performed, first mapping reads to the UniVec database of common laboratory contaminants, then to rRNAs, followed by other small RNA species. Bowtie was used to perform the preprocessing steps (Langmead et al., 2009). MiRNA reads were mapped to human miRNAs annotated in miRbase version 21 (Kozomara et al., 2019). Independent mapping and quantification were performed for mature and precursor miRNAs. Prior to analysis, the raw mature miRNA read counts were normalized and log base 2 transformed using the DESeq2 R package (Love et al., 2014). MiRNAs with a median raw read count of less than five were filtered out.

### Generation of the Longwood paired microRNA and gene expression datasets

We previously reported the Longwood paired miRNA and gene expression datasets, in the context of a larger cohort consisting of 91 samples including 26 pre-chemotherapy diagnostic biopsy specimens paired with 26 post-chemotherapy resection specimens from the same patients, as part of a separate study testing pre chemotherapy biopsy-based miRNA associations with patient survival (Kelly et al., 2013). In our previous publications this dataset is referred to as the “Boston” dataset (Hill et al., 2017; Kelly et al., 2013; Lietz et al., 2020). The Longwood paired dataset presented in the current study is part of the “Boston” dataset, that was not previously analyzed in depth as it relates to dynamic expression changes. All studies were approved by the Beth Israel Deaconess Medical Center and Boston Children’s Hospital IRB, and archival tissue collection was approved by the IRB at both institutions. All experiments were performed in accordance with relevant guidelines and regulations. Detailed methodology is presented in Kelly et al., 2013. In short Illumina cDNA-mediated annealing, selection, extension, and ligation arrays (DASL arrays) generated miRNA profiling data for 91 FFPE samples, and mRNA profiling data for 42 FFPE samples. For pre and post chemotherapy miRNA differential expression testing, miRNAs with expression variance in the bottom tercile across the entire dataset were filtered out. MiRNA and gene expression data for the Longwood miRNA and gene expression datasets are available at the Gene Expression Omnibus under accession GSE39058.

### Public data acquisition and processing

The National Cancer Institute Therapeutically Applicable Research to Generate Effective Treatments (TARGET) initiative’s publicly available osteosarcoma Applied Biosystems MegaPlex TaqMan miRNA and mRNA-seq datasets were downloaded from the TARGET data matrix (https://ocg.cancer.gov/programs/target/data-matrix). At the time data was downloaded, the miRNA and mRNA-seq datasets provided data for 86 and 93 clinically annotated high grade osteosarcoma samples, respectively. Sample selection, experimental, and data processing methodology carried out by the TARGET working group is provided on the TARGET Project Experimental Methods page (https://ocg.cancer.gov/programs/target/target-methods). We applied the 2^-ΔCt^ transformation to the downloaded normalized miRNA data prior to analysis (Schmittgen & Livak, 2008). The gene read count level RNA-seq was further processed by filtering out transcripts less than 200 bases in length and applying *DESeq2* based normalization and log base 2 transformation to the data (Love et al., 2014).

### Differential expression testing and standard statistical tests

*DESeq2* was used to test for differential expression testing or RNA-seq data (Love et al., 2014). A paired analysis design was used for instances where biopsy and resection samples were collected from the same patient.

*Limma* was used for differential expression testing of microarray data (Ritchie et al., 2015). A paired analysis design was again used in instances where biopsy and resection samples were collected from the same patient. Where specified, the false discovery rate (FDR) was calculated using the Benjamini and Hochberg step up procedure (Benjamini & Hochberg, 1995). Pearson’s r statistic were used to test correlations between two variables. The hypergeometric test was used to test enrichment. Association between categorical variables were evaluated with the chi-square test. Hierarchical clustering was performed using the correlation distance and average linkage approaches.

### MiRNA target prediction and pathway enrichment

MiRNA targets were predicted computationally using the DIANA-microTv5.0 server using a microT threshold of 0.7 (Paraskevopoulou et al., 2013). The threshold was tuned to maximize the number of targets per miRNA in the candidate profiles to between 500 and 1500.

Functional annotation of miRNA target genes was performed using DIANA-miRPath v3.0 (Vlachos et al., 2015). Genesets from the Kyoto Encyclopedia of Genes and Genomes (KEGG) and Gene Ontology (GO) databases were analyzed on the DIANA web server using both standard hypergeometric distributions and more stringent unbiased empirical distributions to account for biases encountered by traditional enrichment statistics with miRNA target gene sets (Ashburner et al., 2000; Bleazard et al., 2015; Consortium, 2021; Kanehisa et al., 2023; Kanehisa & Goto, 2000; Paraskevopoulou et al., 2013). MiRPath analysis was run using the meta-analysis pathways union analysis mode for the hypergeometric distribution and empirical distribution tests. The meta-analysis approach pools pathway enrichment from each miRNA’s target set and then provides a single enrichment estimate for the entire miRNA profile, similar to a meta-analysis. Clustered heatmaps depicting the significantly enriched pathways was generated using the MiRPath server.

### Predictive biomarker discovery

The Profiling Relative Inhibition Simultaneously in Mixtures (PRISM) secondary screen with dose-dependent drug response information and the Broad Institute-Novartis Institutes for BioMedical Research Cancer Cell Line Encyclopedia (CCLE) project NanoString miRNA data were downloaded from the Cancer Dependency Map (DepMap) data portal (Barretina et al., 2012; Corsello et al., 2020; Ghandi et al., 2019; Tsherniak et al., 2017; Yu et al., 2016).

Normalized miRNA levels and drug response area under the curve (AUC) values were used for analysis. Prior to modelling response to a given drug, two thirds of available cell lines with drug sensitivity data were randomly assigned to a training set and the other third to a test set, while maintaining proportional tissue type representation in each set. Simple linear regression was performed using the *glmnet* R package using miRNAs to predict drug response AUC values in the training set (Friedman et al., 2010). The resulting models were then tested in the statistically independent test set. The models were also tested in the small number of available osteosarcoma cell lines. These osteosarcoma cell lines were not excluded from the test set as it was theoretically advantageous to use these most representative datapoints in model training. Statistical testing of the models was not performed in this small subset of cell lines for this reason, as well as the small sample size, avoiding circular logic.

Connectivity Map (CMap) analysis using miRNA gene targets was performed on the CLUE.io web server CLUE Query app (Lamb et al., 2006; Subramanian et al., 2005, 2017). The query was performed with the L1000 Touchstone CMap LINCS 2020 dataset and individual query mode.

### Statistical software

Analysis was performed using SPSS version 24, and NCI BRB-ArrayTools v4.6.0 (Simon et al., 2007), CLUE.io web server (Subramanian et al., 2017), DIANA web server (Paraskevopoulou et al., 2013), and R version 3.5.1 and 4.1.0 with packages *amap* (Lucas, 2019), *dendsort* (Sakai et al., 2014), *DESeq2* (Love et al., 2014), *EnhancedVolcano* (Blighe et al., 2021), *ggplot2* (Wickham, 2009), *glmnet* (Friedman et al., 2010), *hmisc* (Harrell & Dupont, 2021), *limma* (Ritchie et al., 2015), *lumi* (Du et al., 2008), *miRBaseConverter* (T. Xu et al., 2018), *pheatmap* (Kolde, 2019), and *RColorBrewer* (Neuwirth, 2014).

## DATA AVAILABILTY

The Longwood dataset has been deposited in the Gene Expression Omnibus and is publicly available under accession GSE39058. The CCLE miRNA and PRISM drug response datasets are available through the Broad Institute DepMap Portal (https://depmap.org/portal/). The results published here are in part based upon data generated by the Therapeutically Applicable Research to Generate Effective Treatments (https://ocg.cancer.gov/programs/target) initiative, phs000218. The data used for this analysis are available at https://portal.gdc.cancer.gov/projects.

## Data Availability

https://www.ncbi.nlm.nih.gov/geo/query/acc.cgi?acc=GSE39058

https://depmap.org/portal/

https://portal.gdc.cancer.gov/projects

## ACKNOWLEDGMENTS

Acknowledgment is expressed towards Cassandra Garbutt, Haotong Wang, Ruoyu Miao, Jason Kim, and Katherine Janeway for assistance with retrieving clinical data, Cassandra Garbutt and Christie Swett for assistance with specimen retrieval, Cassandra Garbutt for performing RNA extraction, Hal Schneider for technical assistance with generating microarray data, Renee Rubio, Brian Lawney and Yaoyu Wang for technical assistance with sequencing, Jeffrey Goldsmith, Vikram Deshpande, and Antonio Perez-Atayde for performing pathology review, and John Quackenbush for assistance with study design.

## REFERENCES

Ashburner, M., Ball, C. A., Blake, J. A., Botstein, D., Butler, H., Cherry, J. M., Davis, A. P., Dolinski, K., Dwight, S. S., Eppig, J. T., Harris, M. A., Hill, D. P., Issel-Tarver, L., Kasarskis, A., Lewis, S., Matese, J. C., Richardson, J. E., Ringwald, M., Rubin, G. M., & Sherlock, G. (2000). Gene Ontology: tool for the unification of biology. Nature Genetics, 25(1), 25–29. 10.1038/75556

Bacci, G., Bertoni, F., Longhi, A., Ferrari, S., Forni, C., Biagini, R., Bacchini, P., Donati, D., Manfrini, M., Bernini, G., & Lari, S. (2003). Neoadjuvant chemotherapy for high-grade central osteosarcoma of the extremity. Histologic response to preoperative chemotherapy correlates with histologic subtype of the tumor. Cancer, 97(12), 3068–3075. 10.1002/cncr.11456

Barretina, J., Caponigro, G., Stransky, N., Venkatesan, K., Margolin, A. A., Kim, S., Wilson, C. J., Lehar, J., Kryukov, G. v, Sonkin, D., Reddy, A., Liu, M., Murray, L., Berger, M. F., Monahan, J. E., Morais, P., Meltzer, J., Korejwa, A., Jane-Valbuena, J., … Garraway, L. A. (2012). The Cancer Cell Line Encyclopedia enables predictive modelling of anticancer drug sensitivity. Nature, 483(7391), 603–607. nature11003 [pii] 10.1038/nature11003

Batlle, E., & Clevers, H. (2017). Cancer stem cells revisited. Nature Medicine, 23(10), 1124–1134. 10.1038/nm.4409

Beird, H. C., Bielack, S. S., Flanagan, A. M., Gill, J., Heymann, D., Janeway, K. A., Livingston, J. A., Roberts, R. D., Strauss, S. J., & Gorlick, R. (2022). Osteosarcoma. Nature Reviews Disease Primers, 8(1), 77. 10.1038/s41572-022-00409-y

Benjamini, Y., & Hochberg, Y. (1995). Controlling the False Discovery Rate: A Practical and Powerful Approach to Multiple Testing. Journal of the Royal Statistical Society. Series B (Methodological), 57(1), 289–300. http://www.jstor.org/stable/2346101

Bertoni, F., & Bacchini, P. (1998). Classification of bone tumors. European Journal of Radiology, 27, S74–S76. 10.1016/S0720-048X(98)00046-1

Bhatia, V., Yadav, A., Tiwari, R., Nigam, S., Goel, S., Carskadon, S., Gupta, N., Goel, A., Palanisamy, N., & Ateeq, B. (2019). Epigenetic Silencing of miRNA-338-5p and miRNA-421 Drives SPINK1-Positive Prostate Cancer. Clinical Cancer Research, 25(9), 2755–2768. 10.1158/1078-0432.CCR-18-3230

Bielack, S. S., Smeland, S., Whelan, J. S., Marina, N., Jovic, G., Hook, J. M., Krailo, M. D., Gebhardt, M., Papai, Z., Meyer, J., Nadel, H., Randall, R. L., Deffenbaugh, C., Nagarajan, R., Brennan, B., Letson, G. D., Teot, L. A., Goorin, A., Baumhoer, D., … investigators, E.-. (2015). Methotrexate, Doxorubicin, and Cisplatin (MAP) Plus Maintenance Pegylated Interferon Alfa-2b Versus MAP Alone in Patients With Resectable High-Grade Osteosarcoma and Good Histologic Response to Preoperative MAP: First Results of the EURAMOS-1 Good Response Randomized Controlled Trial. Journal of Clinical Oncology, 33(20), 2279–2287. 10.1200/JCO.2014.60.0734

Bishop, M. W., Janeway, K. A., & Gorlick, R. (2016). Future directions in the treatment of osteosarcoma. Current Opinions in Pediatrics, 28(1), 26–33. 10.1097/MOP.0000000000000298 [doi]

Bleazard, T., Lamb, J. A., & Griffiths-Jones, S. (2015). Bias in microRNA functional enrichment analysis. Bioinformatics, 31(10), 1592–1598. 10.1093/bioinformatics/btv023

Blighe, K., Rana, S., & Lewis, M. (2021). EnhancedVolcano: publication-ready volcano plots with enhanced colouring and labelling. https://github.com/kevinblighe/EnhancedVolcano

Brown, H. K., Tellez-Gabriel, M., & Heymann, D. (2017). Cancer stem cells in osteosarcoma. Cancer Letters, 386, 189–195. 10.1016/j.canlet.2016.11.019

Calin, G. A., & Croce, C. M. (2006). MicroRNA signatures in human cancers. Nature Reviews Cancer, 6(11), 857– 866. nrc1997 [pii] 10.1038/nrc1997

Calin, G. A., Ferracin, M., Cimmino, A., di Leva, G., Shimizu, M., Wojcik, S. E., Iorio, M. v, Visone, R., Sever, N. I., Fabbri, M., Iuliano, R., Palumbo, T., Pichiorri, F., Roldo, C., Garzon, R., Sevignani, C., Rassenti, L., Alder, H., Volinia, S., … Croce, C. M. (2005). A MicroRNA signature associated with prognosis and progression in chronic lymphocytic leukemia. New England Journal of Medicine, 353(17), 1793–1801. 353/17/1793 [pii] 10.1056/NEJMoa050995

Cao, Z. G., Li, J. J., Yao, L., Huang, Y. N., Liu, Y. R., Hu, X., Song, C. G., & Shao, Z. M. (2016). High expression of microRNA-454 is associated with poor prognosis in triple-negative breast cancer. Oncotarget, 7(40), 64900– 64909.

Carter, L. E., Kilroy, G., Gimble, J. M., & Floyd, Z. E. (2012). An improved method for isolation of RNA from bone. BMC Biotechnology, 12, 5. 1472-6750-12-5 [pii] 10.1186/1472-6750-12-5 [doi]

Chauveinc, L., Mosseri, V., Quintana, E., Desjardins, L., Schlienger, P., Doz, F., & Dutrillaux, B. (2001). Osteosarcoma following retinoblastoma: Age at onset and latency period. Ophthalmic Genetics, 22(2), 77–88. 10.1076/opge.22.2.77.2228

Chen, X., Bahrami, A., Pappo, A., Easton, J., Dalton, J., Hedlund, E., Ellison, D., Shurtleff, S., Wu, G., Wei, L., Parker, M., Rusch, M., Nagahawatte, P., Wu, J., Mao, S., Boggs, K., Mulder, H., Yergeau, D., Lu, C., … st. Jude Childrenâ€TMs Research Hospitalâ€“Washington University Pediatric Cancer Genome, P. (2014). Recurrent somatic structural variations contribute to tumorigenesis in pediatric osteosarcoma. Cell Reports, 7(1), 104– 112. 10.1016/j.celrep.2014.03.003

Chen, Y., Ren, C., Yang, L., Nai, M., Xu, Y., Zhang, F., & Liu, Y. (2019). MicroRNA let 7d 5p rescues ovarian cancer cell apoptosis and restores chemosensitivity by regulating the p53 signaling pathway via HMGA1. International Journal of Oncology, 54(5), 1771–1784.

Cole, S., Gianferante, D. M., Zhu, B., & Mirabello, L. (2022). Osteosarcoma: a Surveillance, Epidemiology, and End Results program-based analysis from 1975 to 2017. Cancer, n/a(n/a). 10.1002/cncr.34163

Consortium, T. G. O. (2021). The Gene Ontology resource: enriching a GOld mine. Nucleic Acids Research, 49(D1), D325–D334. 10.1093/nar/gkaa1113

Corsello, S. M., Bittker, J. A., Liu, Z., Gould, J., McCarren, P., Hirschman, J. E., Johnston, S. E., Vrcic, A., Wong, B., Khan, M., Asiedu, J., Narayan, R., Mader, C. C., Subramanian, A., & Golub, T. R. (2017). The Drug Repurposing Hub: a next-generation drug library and information resource. Nature Medicine, 23(4), 405–408. nm.4306 [pii] 10.1038/nm.4306

Corsello, S. M., Nagari, R. T., Spangler, R. D., Rossen, J., Kocak, M., Bryan, J. G., Humeidi, R., Peck, D., Wu, X., Tang, A. A., Wang, V. M., Bender, S. A., Lemire, E., Narayan, R., Montgomery, P., Ben-David, U., Garvie, C. W., Chen, Y., Rees, M. G., … Golub, T. R. (2020). Discovering the anticancer potential of non-oncology drugs by systematic viability profiling. Nature Cancer, 1(2), 235–248. 10.1038/s43018-019-0018-6

Du, P., Kibbe, W. A., & Lin, S. M. (2008). lumi: a pipeline for processing Illumina microarray. Bioinformatics, 24(13), 1547–1548. btn224 [pii] 10.1093/bioinformatics/btn224

Duan, Z., Gao, Y., Shen, J., Choy, E., Cote, G., Harmon, D., Bernstein, K., Lozano-Calderon, S., Mankin, H., & Hornicek, F. J. (2017). miR-15b modulates multidrug resistance in human osteosarcoma in vitro and in vivo. Molecular Oncology, 11(2), 151–166. 10.1002/1878-0261.12015

Esquela-Kerscher, A., & Slack, F. J. (2006). Oncomirs — microRNAs with a role in cancer. Nature Reviews Cancer, 6(4), 259–269. 10.1038/nrc1840

Fan, J.-B., Yeakley, J. M., Bibikova, M., Chudin, E., Wickham, E., Chen, J., Doucet, D., Rigault, P., Zhang, B., Shen, R., McBride, C., Li, H.-R., Fu, X.-D., Oliphant, A., Barker, D. L., & Chee, M. S. (2004). A Versatile Assay for High-Throughput Gene Expression Profiling on Universal Array Matrices. Genome Research, 14(5), 878–885.

Feng, W., Dean, D. C., Hornicek, F. J., Spentzos, D., Hoffman, R. M., Shi, H., & Duan, Z. (2020). Myc is a prognostic biomarker and potential therapeutic target in osteosarcoma. Therapeutic Advances in Medical Oncology, 12, 1758835920922055–1758835920922055. 10.1177/1758835920922055

Friedman, J. H., Hastie, T., & Tibshirani, R. (2010). Regularization Paths for Generalized Linear Models via Coordinate Descent. Journal of Statistical Software, 33(1), 1–22. 10.18637/jss.v033.i01

German, J. (1997). Bloom’s syndrome. XX. The first 100 cancers. Cancer Genetics and Cytogenetics, 93(1), 100– 106. 10.1016/S0165-4608(96)00336-6

Ghandi, M., Huang, F. W., Jane-Valbuena, J., Kryukov, G. v, Lo, C. C., McDonald 3rd, E. R., Barretina, J., Gelfand, E. T., Bielski, C. M., Li, H., Hu, K., Andreev-Drakhlin, A. Y., Kim, J., Hess, J. M., Haas, B. J., Aguet, F., Weir, B. A., Rothberg, M. v, Paolella, B. R., … Sellers, W. R. (2019). Next-generation characterization of the Cancer Cell Line Encyclopedia. Nature, 569(7757), 503–508. 10.1038/s41586-019-1186-3 [doi] 10.1038/s41586-019-1186-3 [pii]

Gill, J., & Gorlick, R. (2021). Advancing therapy for osteosarcoma. Nature Reviews Clinical Oncology, 18(10), 609–624. 10.1038/s41571-021-00519-8

Glaich, O., Parikh, S., Bell, R. E., Mekahel, K., Donyo, M., Leader, Y., Shayevitch, R., Sheinboim, D., Yannai, S., Hollander, D., Melamed, Z., Lev-Maor, G., Ast, G., & Levy, C. (2019). DNA methylation directs microRNA biogenesis in mammalian cells. Nature Communications, 10(1), 5657. 10.1038/s41467-019-13527-1

Hanahan, D. (2022). Hallmarks of Cancer: New Dimensions. Cancer Discovery, 12(1), 31–46. 10.1158/2159-8290.CD-21-1059

Harrell, F., & Dupont, C. (2021). Harrell Miscellaneous. https://cran.r-project.org/web/packages/Hmisc/index.html

Hill, K. E., Kelly, A. D., Kuijjer, M. L., Barry, W., Rattani, A., Garbutt, C. C., Kissick, H., Janeway, K., Perez-Atayde, A., Goldsmith, J., Gebhardt, M. C., Arredouani, M. S., Cote, G., Hornicek, F., Choy, E., Duan, Z., Quackenbush, J., Haibe-Kains, B., & Spentzos, D. (2017). An imprinted non-coding genomic cluster at 14q32 defines clinically relevant molecular subtypes in osteosarcoma across multiple independent datasets. Journal of Hematology & Oncology, 10(1), 107. 10.1186/s13045-017-0465-4 10.1186/s13045-017-0465-4 [pii]

Hou, N., Han, J., Li, J., Liu, Y., Qin, Y., Ni, L., Song, T., & Huang, C. (2014). MicroRNA Profiling in Human Colon Cancer Cells during 5-Fluorouracil-Induced Autophagy. PLOS ONE, 9(12), e114779-. 10.1371/journal.pone.0114779

Ishikawa, Y., Miller, R. W., Machinami, R., Sugano, H., & Goto, M. (2000). Atypical Osteosarcomas in Werner Syndrome (Adult Progeria). Japanese Journal of Cancer Research, 91(12), 1345–1349. 10.1111/j.1349-7006.2000.tb00924.x

Ji, D., Zhan, T., Li, M., Yao, Y., Jia, J., Yi, H., Qiao, M., Xia, J., Zhang, Z., Ding, H., Song, C., Han, Y., & Gu, J. (2018). Enhancement of Sensitivity to Chemo/Radiation Therapy by Using miR-15b against DCLK1 in Colorectal Cancer. Stem Cell Reports, 11(6), 1506–1522. 10.1016/j.stemcr.2018.10.015

Jin, Y., Wei, J., Xu, S., Guan, F., Yin, L., & Zhu, H. (2019). miR 210 3p regulates cell growth and affects cisplatin sensitivity in human ovarian cancer cells via targeting E2F3. Molecular Medicine Reports, 19(6), 4946–4954.

Johnstone, S. E., Reyes, A., Qi, Y., Adriaens, C., Hegazi, E., Pelka, K., Chen, J. H., Zou, L. S., Drier, Y., Hecht, V., Shoresh, N., Selig, M. K., Lareau, C. A., Iyer, S., Nguyen, S. C., Joyce, E. F., Hacohen, N., Irizarry, R. A., Zhang, B., … Bernstein, B. E. (2020). Large-Scale Topological Changes Restrain Malignant Progression in Colorectal Cancer. Cell, 182(6), 1474–1489.e23. 10.1016/j.cell.2020.07.030

Jonas, S., & Izaurralde, E. (2015). Towards a molecular understanding of microRNA-mediated gene silencing. Nature Reviews Genetics, 16(7), 421–433. 10.1038/nrg3965

Kanehisa, M., Furumichi, M., Sato, Y., Kawashima, M., & Ishiguro-Watanabe, M. (2023). KEGG for taxonomy-based analysis of pathways and genomes. Nucleic Acids Research, 51(D1), D587–D592. 10.1093/nar/gkac963

Kanehisa, M., & Goto, S. (2000). KEGG: Kyoto Encyclopedia of Genes and Genomes. Nucleic Acids Research, 28(1), 27–30. 10.1093/nar/28.1.27

Kanehisa, M., Sato, Y., Furumichi, M., Morishima, K., & Tanabe, M. (2019). New approach for understanding genome variations in KEGG. Nucleic Acids Research, 47(D1), D590–D595. 5128935 [pii] 10.1093/nar/gky962 [doi]

Kelly, A. D., Haibe-Kains, B., Janeway, K. A., Hill, K. E., Howe, E., Goldsmith, J., Kurek, K., Perez-Atayde, A. R., Francoeur, N., Fan, J. B., April, C., Schneider, H., Gebhardt, M. C., Culhane, A., Quackenbush, J., & Spentzos, D. (2013). MicroRNA paraffin-based studies in osteosarcoma reveal reproducible independent prognostic profiles at 14q32. Genome Medicine, 5(1), 2. gm406 [pii] 10.1186/gm406

Kolde, R. (2019). Pretty Heatmaps. https://cran.r-project.org/web/packages/pheatmap/index.html

Kozomara, A., Birgaoanu, M., & Griffiths-Jones, S. (2019). miRBase: from microRNA sequences to function. Nucleic Acids Research, 47(D1), D155–D162. 5179337 [pii] 10.1093/nar/gky1141 [doi]

Lai, J., Yang, H., Zhu, Y., Ruan, M., Huang, Y., & Zhang, Q. (2019). MiR-7-5p-mediated downregulation of PARP1 impacts DNA homologous recombination repair and resistance to doxorubicin in small cell lung cancer. BMC Cancer, 19(1), 602. 10.1186/s12885-019-5798-7

Lamb, J., Crawford, E. D., Peck, D., Modell, J. W., Blat, I. C., Wrobel, M. J., Lerner, J., Brunet, J.-P., Subramanian, A., Ross, K. N., Reich, M., Hieronymus, H., Wei, G., Armstrong, S. A., Haggarty, S. J., Clemons, P. A., Wei, R., Carr, S. A., Lander, E. S., & Golub, T. R. (2006). The Connectivity Map: Using Gene-Expression Signatures to Connect Small Molecules, Genes, and Disease. Science, 313(5795), 1929–1935. 10.1126/science.1132939

Langmead, B., Trapnell, C., Pop, M., & Salzberg, S. L. (2009). Ultrafast and memory-efficient alignment of short DNA sequences to the human genome. Genome Biology, 10(3), R25. gb-2009-10-3-r25 [pii] 10.1186/gb-2009-10-3-r25

Larizza, L., Roversi, G., & Volpi, L. (2010). Rothmund-Thomson syndrome. Orphanet Journal of Rare Diseases, 5(1), 2. 10.1186/1750-1172-5-2

Li, F. P., Fraumeni Jr., J. F., Mulvihill, J. J., Blattner, W. A., Dreyfus, M. G., Tucker, M. A., & Miller, R. W. (1988). A Cancer Family Syndrome in Twenty-four Kindreds1. Cancer Research, 48(18), 5358–5362.

Li, H.-Y., Liang, J.-L., Kuo, Y.-L., Lee, H.-H., Calkins, M. J., Chang, H.-T., Lin, F.-C., Chen, Y.-C., Hsu, T.-I., Hsiao, M., Ger, L.-P., & Lu, P.-J. (2017). miR-105/93-3p promotes chemoresistance and circulating miR-105/93-3p acts as a diagnostic biomarker for triple negative breast cancer. Breast Cancer Research, 19(1), 133. 10.1186/s13058-017-0918-2

Lietz, C. E., Garbutt, C., Barry, W. T., Deshpande, V., Chen, Y.-L., Lozano-Calderon, S. A., Wang, Y., Lawney, B., Ebb, D., Cote, G. M., Duan, Z., Hornicek, F. J., Choy, E., Nielsen, G. P., Haibe-Kains, B., Quackenbush, J., & Spentzos, D. (2020). MicroRNA-mRNA networks define translatable molecular outcome phenotypes in osteosarcoma. Scientific Reports, 10(4409). 10.1038/s41598-020-61236-3

Lietz, C. E., Newman, E. T., Kelly, A. D., Xiang, D. H., Zhang, Z., Luscko, C. A., Lozano-Calderon, S. A., Ebb, D. H., Raskin, K. A., Cote, G. M., Choy, E., Nielsen, G. P., Haibe-Kains, B., Aryee, M. J., & Spentzos, D. (2022). Genome- wide DNA methylation patterns reveal clinically relevant predictive and prognostic subtypes in human osteosarcoma. Communications Biology, 5(1), 213. 10.1038/s42003-022-03117-1

Lin, S. M., Du, P., Huber, W., & Kibbe, W. A. (2008). Model-based variance-stabilizing transformation for Illumina microarray data. Nucleic Acids Research, 36(2), e11. gkm1075 [pii] 10.1093/nar/gkm1075

Lipton, J. M., Federman, N., Khabbaze, Y., Schwartz, C. L., Hilliard, L. M., Clark, J. I., & Vlachos, A. (2001). Osteogenic Sarcoma Associated With Diamond[ndash]Blackfan Anemia[colon] A Report From the Diamond[ndash]Blackfan Anemia Registry. Journal of Pediatric Hematology/Oncology, 23(1). https://journals.lww.com/jpho-online/Fulltext/2001/01000/Osteogenic_Sarcoma_Associated_With.9.aspx

Liu, J., Valencia-Sanchez, M. A., Hannon, G. J., & Parker, R. (2005). MicroRNA-dependent localization of targeted mRNAs to mammalian P-bodies. Nature Cell Biology, 7(7), 719–723. 10.1038/ncb1274

Livak, K. J., & Schmittgen, T. D. (2001). Analysis of relative gene expression data using real-time quantitative PCR and the 2(-Delta Delta C(T)) Method. Methods, 25(4), 402–408. 10.1006/meth.2001.1262 [doi] S1046-2023(01)91262-9 [pii]

Lorenz, S., Baroy, T., Sun, J., Nome, T., Vodak, D., Bryne, J. C., Hakelien, A. M., Fernandez-Cuesta, L., Mohlendick, B., Rieder, H., Szuhai, K., Zaikova, O., Ahlquist, T. C., Thomassen, G. O., Skotheim, R. I., Lothe, R. A., Tarpey, P. S., Campbell, P., Flanagan, A., … Meza-Zepeda, L. A. (2016). Unscrambling the genomic chaos of osteosarcoma reveals extensive transcript fusion, recurrent rearrangements and frequent novel TP53 aberrations. Oncotarget, 7(5), 5273–5288. 6567 [pii] 10.18632/oncotarget.6567 [doi]

Love, M. I., Huber, W., & Anders, S. (2014). Moderated estimation of fold change and dispersion for RNA-seq data with DESeq2. Genome Biology, 15(12), 550. s13059-014-0550-8 [pii] 10.1186/s13059-014-0550-8

Lozano Calderon, S. A., Garbutt, C., Kim, J., Lietz, C. E., Chen, Y. L., Bernstein, K., Chebib, I., Nielsen, G. P., Deshpande, V., Rubio, R., Wang, Y. E., Quackenbush, J., Delaney, T., Raskin, K., Schwab, J., Cote, G., & Spentzos, D. (2019). Clinical and Molecular Analysis of Pathologic Fracture-associated Osteosarcoma: MicroRNA profile Is Different and Correlates with Prognosis. Clinical Orthopaedics and Related Research, 477(9), 2114–2126. 10.1097/CORR.0000000000000867 [doi]

Lu, J., Getz, G., Miska, E. A., Alvarez-Saavedra, E., Lamb, J., Peck, D., Sweet-Cordero, A., Ebert, B. L., Mak, R. H., Ferrando, A. A., Downing, J. R., Jacks, T., Horvitz, H. R., & Golub, T. R. (2005). MicroRNA expression profiles classify human cancers. Nature, 435(7043), 834–838. nature03702 [pii] 10.1038/nature03702

Lu, M., Wang, C., Chen, W., Mao, C., & Wang, J. (2018). miR-654-5p Targets GRAP to Promote Proliferation, Metastasis, and Chemoresistance of Oral Squamous Cell Carcinoma Through Ras/MAPK Signaling. DNA and Cell Biology, 37(4), 381–388. 10.1089/dna.2017.4095

Lucas, A. (2019). Another Multidimensional Analysis Package. https://cran.r-project.org/web/packages/amap/index.html

Marina, N. M., Smeland, S., Bielack, S. S., Bernstein, M., Jovic, G., Krailo, M. D., Hook, J. M., Arndt, C., van den Berg, H., Brennan, B., Brichard, B., Brown, K. L., Butterfass-Bahloul, T., Calaminus, G., Daldrup-Link, H. E., Eriksson, M., Gebhardt, M. C., Gelderblom, H., Gerss, J., … Whelan, J. S. (2016). Comparison of MAPIE versus MAP in patients with a poor response to preoperative chemotherapy for newly diagnosed high-grade osteosarcoma (EURAMOS-1): an open-label, international, randomised controlled trial. Lancet Oncology, 17(10), 1396–1408. S1470-2045(16)30214-5 [pii] 10.1016/S1470-2045(16)30214-5

Meltzer, P. S., & Helman, L. J. (2021). New Horizons in the Treatment of Osteosarcoma. New England Journal of Medicine, 385(22), 2066–2076. 10.1056/NEJMra2103423

Mi, S., Lu, J., Sun, M., Li, Z., Zhang, H., Neilly, M. B., Wang, Y., Qian, Z., Jin, J., Zhang, Y., Bohlander, S. K., le Beau, M. M., Larson, R. A., Golub, T. R., Rowley, J. D., & Chen, J. (2007). MicroRNA expression signatures accurately discriminate acute lymphoblastic leukemia from acute myeloid leukemia. Proceedings of the National Academy of Sciences, 104(50), 19971–19976. 0709313104 [pii] 10.1073/pnas.0709313104

Mirabello, L., Troisi, R. J., & Savage, S. A. (2009). Osteosarcoma incidence and survival rates from 1973 to 2004: data from the Surveillance, Epidemiology, and End Results Program. Cancer, 115(7), 1531–1543. 10.1002/cncr.24121

Mirabello, L., Zhu, B., Koster, R., Karlins, E., Dean, M., Yeager, M., Gianferante, M., Spector, L. G., Morton, L. M., Karyadi, D., Robison, L. L., Armstrong, G. T., Bhatia, S., Song, L., Pankratz, N., Pinheiro, M., Gastier-Foster, J. M., Gorlick, R., de Toledo, S. R. C., … Savage, S. A. (2020). Frequency of Pathogenic Germline Variants in Cancer- Susceptibility Genes in Patients With Osteosarcoma. JAMA Oncology, 6(5), 724–734. 2762589 [pii] coi200005 [pii] 10.1001/jamaoncol.2020.0197 [doi]

Neuwirth, E. (2014). ColorBrewer Palettes. https://cran.r-project.org/web/packages/RColorBrewer/index.html

Niu, Y., Tong, J., Shi, X., & Zhang, T. (2020). MicroRNA 654 3p enhances cisplatin sensitivity by targeting QPRT and inhibiting the PI3K/AKT signaling pathway in ovarian cancer cells. Experimental and Therapeutic Medicine, 20(2), 1467–1479.

Ooi, C. H., Oh, H. K., Wang, H. Z., Tan, A. L., Wu, J., Lee, M., Rha, S. Y., Chung, H. C., Virshup, D. M., & Tan, P. (2011). A densely interconnected genome-wide network of microRNAs and oncogenic pathways revealed using gene expression signatures. PLoS Genetics, 7(12), e1002415. 10.1371/journal.pgen.1002415 PGENETICS-D-11-00909 [pii]

Paik, S., Shak, S., Tang, G., Kim, C., Baker, J., Cronin, M., Baehner, F. L., Walker, M. G., Watson, D., Park, T., Hiller, W., Fisher, E. R., Wickerham, D. L., Bryant, J., & Wolmark, N. (2004). A multigene assay to predict recurrence of tamoxifen-treated, node-negative breast cancer. New England Journal of Medicine, 351(27), 2817–2826. NEJMoa041588 [pii] 10.1056/NEJMoa041588

Paraskevopoulou, M. D., Georgakilas, G., Kostoulas, N., Vlachos, I. S., Vergoulis, T., Reczko, M., Filippidis, C., Dalamagas, T., & Hatzigeorgiou, A. G. (2013). DIANA-microT web server v5.0: service integration into miRNA functional analysis workflows. Nucleic Acids Research, 41(W1), W169–W173. 10.1093/nar/gkt393

Perry, J. A., Kiezun, A., Tonzi, P., van Allen, E. M., Carter, S. L., Baca, S. C., Cowley, G. S., Bhatt, A. S., Rheinbay, E., Pedamallu, C. S., Helman, E., Taylor-Weiner, A., McKenna, A., DeLuca, D. S., Lawrence, M. S., Ambrogio, L., Sougnez, C., Sivachenko, A., Walensky, L. D., … Janeway, K. A. (2014). Complementary genomic approaches highlight the PI3K/mTOR pathway as a common vulnerability in osteosarcoma. Proceedings of the National Academy of Sciences, 111(51), E5564–73. 10.1073/pnas.1419260111

Picci, P., Sangiorgi, L., Rougraff, B. T., Neff, J. R., Casadei, R., & Campanacci, M. (1994). Relationship of chemotherapy-induced necrosis and surgical margins to local recurrence in osteosarcoma. Journal of Clinical Oncology, 12(12), 2699–2705. 10.1200/JCO.1994.12.12.2699

Ritchie, M. E., Phipson, B., Wu, D., Hu, Y., Law, C. W., Shi, W., & Smyth, G. K. (2015). limma powers differential expression analyses for RNA-sequencing and microarray studies. Nucleic Acids Research, 43(7), e47–e47. 10.1093/nar/gkv007

Roberts, R. D., Lizardo, M. M., Reed, D. R., Hingorani, P., Glover, J., Allen-Rhoades, W., Fan, T., Khanna, C., Sweet-Cordero, E. A., Cash, T., Bishop, M. W., Hegde, M., Sertil, A. R., Koelsche, C., Mirabello, L., Malkin, D., Sorensen, P. H., Meltzer, P. S., Janeway, K. A., … Crompton, B. D. (2019). Provocative questions in osteosarcoma basic and translational biology: A report from the Children’s Oncology Group. Cancer, 125(20), 3514–3525. 10.1002/cncr.32351 [doi]

Rosen, G., Caparros, B., Huvos, A. G., Kosloff, C., Nirenberg, A., Cacavio, A., Marcove, R. C., Lane, J. M., Mehta, B., & Urban, C. (1982). Preoperative chemotherapy for osteogenic sarcoma: Selection of postoperative adjuvant chemotherapy based on the response of the primary tumor to preoperative chemotherapy. Cancer, 49(6), 1221–1230. 10.1002/1097-0142(19820315)49:6<1221::AID-CNCR2820490625>3.0.CO;2-E

Rosen, G., Marcove, R. C., Caparros, B., Nirenberg, A., Kosloff, C., & Huvos, A. G. (1979). Primary osteogenic sarcoma: the rationale for preoperative chemotherapy and delayed surgery. Cancer, 43(6), 2163–2177. 10.1002/1097-0142(197906)43:6<2163::aid-cncr2820430602>3.0.co;2-s [doi]

Rosen, G., Murphy, M. L., Huvos, A. G., Gutierrez, M., & Marcove, R. C. (1976). Chemotherapy, en bloc resection, and prosthetic bone replacement in the treatment of osteogenic sarcoma. Cancer, 37(1), 1–11. 10.1002/1097-0142(197601)37:1<1::AID-CNCR2820370102>3.0.CO;2-3

Rueda, A., Barturen, G., Lebron, R., Gomez-Martin, C., Alganza, A., Oliver, J. L., & Hackenberg, M. (2015). sRNAtoolbox: an integrated collection of small RNA research tools. Nucleic Acids Research, 43(W1), W467–73. gkv555 [pii] 10.1093/nar/gkv555 [doi]

Sakai, R., Winand, R., Verbeiren, T., Moere, A. v, & Aerts, J. (2014). dendsort: modular leaf ordering methods for dendrogram representations in R. F1000Research, 3, 177. 10.12688/f1000research.4784.1 [doi]

Sarver, A. L., Thayanithy, V., Scott, M. C., Cleton-Jansen, A. M., Hogendoorn, P. C., Modiano, J. F., & Subramanian, S. (2013). MicroRNAs at the human 14q32 locus have prognostic significance in osteosarcoma. Orphanet Journal of Rare Diseases, 8, 7. 1750-1172-8-7 [pii] 10.1186/1750-1172-8-7

Sayles, L. C., Breese, M. R., Koehne, A. L., Leung, S. G., Lee, A. G., Liu, H. Y., Spillinger, A., Shah, A. T., Tanasa, B., Straessler, K., Hazard, F. K., Spunt, S. L., Marina, N., Kim, G. E., Cho, S. J., Avedian, R. S., Mohler, D. G., Kim, M. O., DuBois, S. G., … Sweet-Cordero, E. A. (2019). Genome-Informed Targeted Therapy for Osteosarcoma. Cancer Discovery, 9(1), 46–63. 2159-8290.CD-17-1152 [pii] 10.1158/2159-8290.CD-17-1152 [doi]

Schmittgen, T. D., & Livak, K. J. (2008). Analyzing real-time PCR data by the comparative C(T) method. Nature Protocols, 3(6), 1101–1108. 10.1038/nprot.2008.73 [doi]

Schreiber, R., Mezencev, R., Matyunina, L. v, & McDonald, J. F. (2016). Evidence for the role of microRNA 374b in acquired cisplatin resistance in pancreatic cancer cells. Cancer Gene Therapy, 23(8), 241–245. 10.1038/cgt.2016.23

Siitonen, H. A., Sotkasiira, J., Biervliet, M., Benmansour, A., Capri, Y., Cormier-Daire, V., Crandall, B., Hannula-Jouppi, K., Hennekam, R., Herzog, D., Keymolen, K., Lipsanen-Nyman, M., Miny, P., Plon, S. E., Riedl, S., Sarkar, A., Vargas, F. R., Verloes, A., Wang, L. L., … Kestilä, M. (2009). The mutation spectrum in RECQL4 diseases. European Journal of Human Genetics, 17(2), 151–158. 10.1038/ejhg.2008.154

Simon, R., Lam, A., Li, M. C., Ngan, M., Menenzes, S., & Zhao, Y. (2007). Analysis of gene expression data using BRB-ArrayTools. Cancer Informatics, 3, 11–17.

Subramanian, A., Narayan, R., Corsello, S. M., Peck, D. D., Natoli, T. E., Lu, X., Gould, J., Davis, J. F., Tubelli, A. A., Asiedu, J. K., Lahr, D. L., Hirschman, J. E., Liu, Z., Donahue, M., Julian, B., Khan, M., Wadden, D., Smith, I. C., Lam, D., … Golub, T. R. (2017). A Next Generation Connectivity Map: L1000 Platform and the First 1,000,000 Profiles. Cell, 171(6), 1437–1452 e17. S0092-8674(17)31309-0 [pii] 10.1016/j.cell.2017.10.049

Subramanian, A., Tamayo, P., Mootha, V. K., Mukherjee, S., Ebert, B. L., Gillette, M. A., Paulovich, A., Pomeroy, S. L., Golub, T. R., Lander, E. S., & Mesirov, J. P. (2005). Gene set enrichment analysis: A knowledge-based approach for interpreting genome-wide expression profiles. Proceedings of the National Academy of Sciences, 102(43), 15545–15550. 10.1073/pnas.0506580102

Tie, Y., Chen, C., Qian, Z., Yuan, H., Wang, H., Tang, H., Peng, Y., Du, X., & Liu, B. (2018). Upregulation of let 7f 5p promotes chemotherapeutic resistance in colorectal cancer by directly repressing several pro apoptotic proteins. Oncology Letters, 15(6), 8695–8702.

Tsherniak, A., Vazquez, F., Montgomery, P. G., Weir, B. A., Kryukov, G., Cowley, G. S., Gill, S., Harrington, W. F., Pantel, S., Krill-Burger, J. M., Meyers, R. M., Ali, L., Goodale, A., Lee, Y., Jiang, G., Hsiao, J., Gerath, W. F. J., Howell, S., Merkel, E., … Hahn, W. C. (2017). Defining a Cancer Dependency Map. Cell, 170(3), 564–576.e16. 10.1016/j.cell.2017.06.010

Vlachos, I. S., Zagganas, K., Paraskevopoulou, M. D., Georgakilas, G., Karagkouni, D., Vergoulis, T., Dalamagas, T., & Hatzigeorgiou, A. G. (2015). DIANA-miRPath v3.0: deciphering microRNA function with experimental support. Nucleic Acids Research, 43(W1), W460–W466. 10.1093/nar/gkv403

Wang, Y., Bao, W., Liu, Y., Wang, S., Xu, S., Li, X., Li, Y., & Wu, S. (2018). miR-98-5p contributes to cisplatin resistance in epithelial ovarian cancer by suppressing miR-152 biogenesis via targeting Dicer1. Cell Death & Disease, 9(5), 447. 10.1038/s41419-018-0390-7

Whelan, J. S., Bielack, S. S., Marina, N., Smeland, S., Jovic, G., Hook, J. M., Krailo, M., Anninga, J., Butterfass- Bahloul, T., Bohling, T., Calaminus, G., Capra, M., Deffenbaugh, C., Dhooge, C., Eriksson, M., Flanagan, A. M., Gelderblom, H., Goorin, A., Gorlick, R., … collaborators, E. (2015). EURAMOS-1, an international randomised study for osteosarcoma: results from pre-randomisation treatment. Annals of Oncology, 26(2), 407–414. 10.1093/annonc/mdu526

Whittle, S. B., Offer, K., Roberts, R. D., LeBlanc, A., London, C., Majzner, R. G., Huang, A. Y., Houghton, P., Alejandro Sweet Cordero, E., Grohar, P. J., Isakoff, M., Bishop, M. W., Stewart, E., Slotkin, E. K., Greengard, E., Borinstein, S. C., Navid, F., Gorlick, R., Janeway, K. A., … Hingorani, P. (2021). Charting a path for prioritization of novel agents for clinical trials in osteosarcoma: A report from the Children’s Oncology Group New Agents for Osteosarcoma Task Force. Pediatric Blood & Cancer, e29188. 10.1002/pbc.29188 [doi]

Wickham, H. (2009). ggplot2: Elegant Graphics for Data Analysis. Springer-Verlag.

Xu, M., Xiao, J., Chen, M., Yuan, L., Li, J., Shen, H., & Yao, S. (2018). miR 149 5p promotes chemotherapeutic resistance in ovarian cancer via the inactivation of the Hippo signaling pathway. International Journal of Oncology, 52(3), 815–827.

Xu, T., Su, N., Liu, L., Zhang, J., Wang, H., Zhang, W., Gui, J., Yu, K., Li, J., & Le, T. D. (2018). miRBaseConverter: an R/Bioconductor package for converting and retrieving miRNA name, accession, sequence and family information in different versions of miRBase. BMC Bioinformatics, 19(19), 514. 10.1186/s12859-018-2531-5

Yamaguchi, T., Toguchida, J., Yamamuro, T., Kotoura, Y., Takada, N., Kawaguchi, N., Kaneko, Y., Nakamura, Y., Sasaki, M. S., & Ishizaki, K. (1992). Allelotype Analysis in Osteosarcomas: Frequent Allele Loss on 3q, 13q, 17p, and 18q. Cancer Research, 52(9), 2419. http://cancerres.aacrjournals.org/content/52/9/2419.abstract

Yu, C., Mannan, A. M., Yvone, G. M., Ross, K. N., Zhang, Y.-L., Marton, M. A., Taylor, B. R., Crenshaw, A., Gould, J. Z., Tamayo, P., Weir, B. A., Tsherniak, A., Wong, B., Garraway, L. A., Shamji, A. F., Palmer, M. A., Foley, M. A., Winckler, W., Schreiber, S. L., … Golub, T. R. (2016). High-throughput identification of genotype-specific cancer vulnerabilities in mixtures of barcoded tumor cell lines. Nature Biotechnology, 34(4), 419–423. 10.1038/nbt.3460

Zhang, Q., Zhang, B., Sun, L., Yan, Q., Zhang, Y., Zhang, Z., Su, Y., & Wang, C. (2018). MicroRNA-130b targets PTEN to induce resistance to cisplatin in lung cancer cells by activating Wnt/β-catenin pathway. Cell Biochemistry and Function, 36(4), 194–202. 10.1002/cbf.3331

Zhao, C., Zhao, Q., Zhang, C., Wang, G., Yao, Y., Huang, X., Zhan, F., Zhu, Y., Shi, J., Chen, J., Yan, F., & Zhang, Y. (2017). miR-15b-5p resensitizes colon cancer cells to 5-fluorouracil by promoting apoptosis via the NF-κB/XIAP axis. Scientific Reports, 7(1), 4194. 10.1038/s41598-017-04172-z

Zhou, Y., Lu, Q., Xu, J., Yan, R., Zhu, J., Xu, J., Jiang, X., Li, J., & Wu, F. (2017). The effect of pathological fractures on the prognosis of patients with osteosarcoma: a meta-analysis of 14 studies. Oncotarget, 8(42), 73037– 73049.

Zhu, H., Yang, J., & Yang, S. (2020). MicroRNA 103a 3p potentiates chemoresistance to cisplatin in non small cell lung carcinoma by targeting neurofibromatosis 1. Experimental and Therapeutic Medicine, 19(3), 1797– 1805.

Ziller, M. J., Gu, H., Müller, F., Donaghey, J., Tsai, L. T.-Y., Kohlbacher, O., de Jager, P. L., Rosen, E. D., Bennett, D. A., Bernstein, B. E., Gnirke, A., & Meissner, A. (2013). Charting a dynamic DNA methylation landscape of the human genome. Nature, 500(7463), 477–481. 10.1038/nature12433

