## Supplementary Table 2 for "A dynamic microRNA profile that tracks a chemotherapy resistance phenotype in osteosarcoma. Implications for novel therapeutics"

### Supplementary Materials

**Supplementary Table 1.** Demographic and clinical characteristics of patients included in the MGH and Longwood cohorts.

| Sample | Dataset | miRNA | mRNA-biopsy | mRNA-resection | Chemo-resistant | Sex | Necrosis (%) | Recurrence | Recurrence Free Survival (months) | Death | Survival Time (months) |
|---|---|---|---|---|---|---|---|---|---|---|---|
| A | MGH | yes | no | no | yes | M | 75 | no | 133 | no | 133 |
| E | MGH | yes | no | no | yes | F | 85 | yes | 3 | yes | 22 |
| I | MGH | yes | no | no | yes | M | 75 | yes | 15 | yes | 69 |
| L | MGH | yes | no | no | yes | M | 85 | no | 143 | no | 143 |
| R | MGH | yes | no | no | yes | M | 50 | yes | 18 | yes | 25 |
| U | MGH | yes | no | no | yes | M | 0 | yes | 61 | yes | 62 |
| V | MGH | yes | no | no | yes | M | 60 | yes | 11 | yes | 59 |
| B | MGH | yes | no | no | no | M | NA | no | 207 | no | 207 |
| G | MGH | yes | no | no | no | M | NA | no | 76 | no | 76 |
| J | MGH | yes | no | no | no | M | NA | yes | 10 | no | 162 |
| K | MGH | yes | no | no | no | M | NA | no | 72 | no | 72 |
| M | MGH | yes | no | no | no | M | NA | no | 78 | no | 78 |
| N | MGH | yes | no | no | no | M | NA | no | 63 | no | 63 |
| O | MGH | yes | no | no | no | F | NA | yes | 16 | yes | 16 |
| 1 | Longwood | yes | yes | no | yes | M | 60 | yes | NA | yes | NA |
| 2 | Longwood | yes | yes | no | yes | M | 65 | no | 28 | no | 28 |
| 3 | Longwood | yes | yes | no | yes | M | 50 | yes | NA | yes | NA |
| 4 | Longwood | yes | yes | yes | yes | F | 15 | no | 34 | no | 34 |
| 5 | Longwood | yes | yes | no | yes | F | 70 | yes | 9 | no | 12 |
| 6 | Longwood | yes | yes | no | yes | F | 80 | no | 50 | no | 50 |
| 7 | Longwood | yes | yes | no | yes | M | 70 | yes | 13 | yes | 40 |
| 8 | Longwood | yes | no | no | yes | F | 30 | no | 66 | no | 66 |
| 9 | Longwood | yes | yes | no | yes | F | 40 | yes | 34 | no | 136 |
| 10 | Longwood | yes | yes | no | yes | F | 30 | no | 124 | no | 124 |
| 11 | Longwood | yes | yes | no | yes | F | 40 | yes | 59 | no | 77 |
| 12 | Longwood | yes | yes | no | yes | F | 65 | no | 50 | no | 50 |
| 13 | Longwood | yes | yes | no | yes | F | 80 | yes | 65 | no | 75 |
| 14 | Longwood | yes | yes | yes | yes | F | 50 | yes | 9 | yes | 13 |
| 15 | Longwood | yes | yes | no | yes | M | 20 | yes | 14 | yes | 23 |
| 16 | Longwood | yes | yes | no | yes | F | 50 | yes | 14 | yes | 20 |
| 17 | Longwood | yes | yes | no | yes | M | 20 | yes | 28 | no | 33 |
| 18 | Longwood | yes | yes | no | yes | M | 50 | no | 0 | no | 0 |
| 19 | Longwood | yes | yes | no | yes | M | 77.5 | yes | 14 | no | 69 |
| 20 | Longwood | yes | yes | yes | yes | M | 40 | no | 6 | no | 6 |
| 21 | Longwood | yes | no | no | yes | F | 80 | yes | 26 | yes | 26 |
| 22 | Longwood | yes | yes | yes | yes | F | 5 | yes | 25 | yes | 25 |
| 23 | Longwood | yes | no | no | yes | F | 22.5 | no | 59 | no | 59 |
| 24 | Longwood | yes | no | no | yes | F | 50 | yes | 11 | no | 25 |
| 25 | Longwood | yes | yes | no | yes | M | 20 | yes | 3 | yes | 6 |
| 26 | Longwood | yes | yes | yes | yes | M | 50 | no | 32 | no | 32 |

**Supplementary Table 2.** CDCRP miRNA – gene target Pearson correlations in the TARGET and Longwood datasets. Included as a separate file.

**Supplementary Figure 1.** CDCRP expression profile in the MGH untreated pairs. Hierarchical clustering of the MGH untreated cohort using expression levels of the CDCRP miRNAs. Note that the CDCRP profile was not expected to differentiate between theses untreated samples.

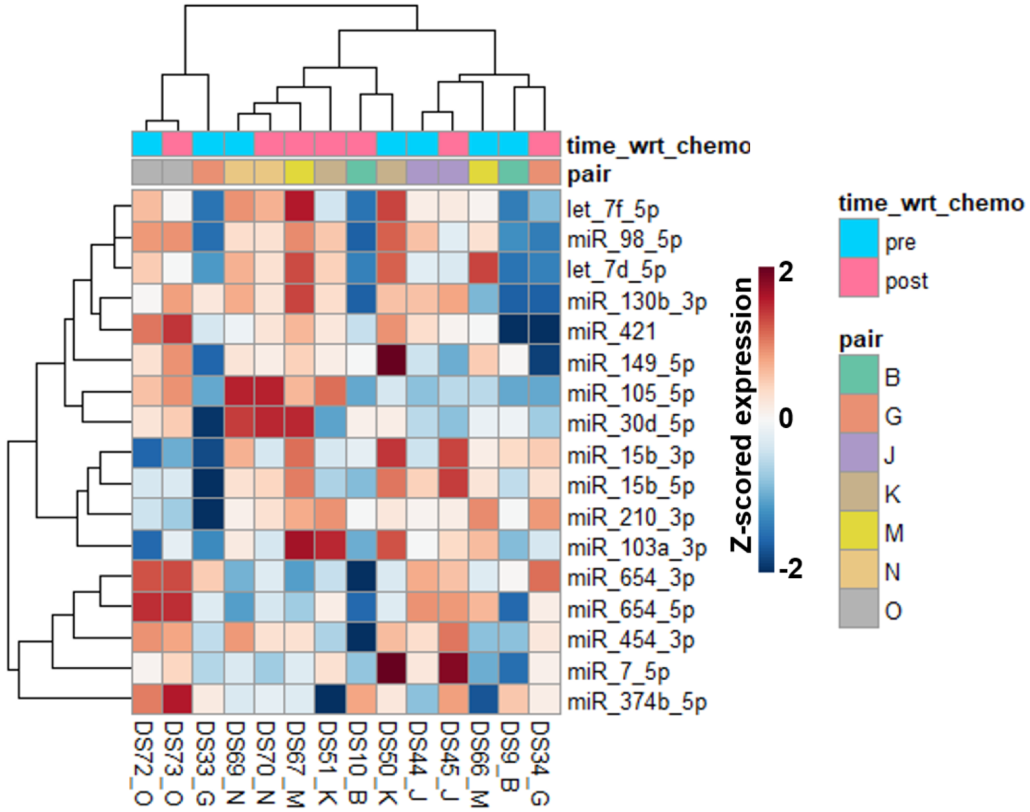
